# Going Digital or Stayin’ Clinical?: Modelling Primary Health Care Consultation Types Using Register Data from Sweden

**DOI:** 10.1101/2024.12.13.24318993

**Authors:** Björn Ekman

## Abstract

**Background:** Primary health care clinics use several different types of consultations, including on-site visits and remote contacts to see patients. However, it is not clear what factors determine the choice of consultation type taking into consideration the full spectrum of options.

**Objectives:** This study uses national register data from Sweden on all primary care consultations in the years 2019 to 2022 (N = 15,3 – 19,5 million) to model four types of consultations: office visits, digital contacts (text messages or video), home visits, and telephone/letter contacts.

**Methods:** The modelling of consultation type takes the cross-sectional, panel, and multilevel structure of the data into consideration fitting multinomial logistic models while controlling for patient, clinic, and context factors.

**Results:** In the current context, the choice of consultation type is determined by both patient (age, socioeconomic status, diagnosis) and clinic (public or private) factors, as well as context factors (reimbursement mechanism). From a policy perspective, only weak evidence is found to suggest that paying clinics for digital contacts increases those types of consultations.

**Conclusions:** The findings may inform the development and implementation of policies and regulations to improve the delivery of primary health care in similar contexts.

## Introduction

Primary health care consultations can take one of several alternative forms. Most broadly, consultations are either in-person or remote. In-person consultations take place either at the clinic or in the patient’s home. Examples of remote consultations include letter and telephone contacts. More recently, remote consultations can be done by means of digital information and communication technologies for health (HIT), such as chat (asynchronous text messages) and video (synchronous image-based communication). While the most common type of consultation is the in-person clinic visit, consultations by means of some type of HIT are increasing. Indeed, during the Covid-19 pandemic, remote consultations by means of both telephone and digital technologies increased in most European and other high-income countries (Fahy and Williams 2021).

While the type of consultation in any particular case is the result of factors on both the patient and the provider side, little is currently known about the determinants of the type of primary care consultation on the level of the overall system, i.e., considering the full range of consultation types. The current evidence on the use of different consultation types has largely focused on some particular type of consultation, such as clinic visits (Blumenfeld and Yaphe 2007; Toal-Sullivan, Dahrouge et al. 2024), digital care (Stoffers 2018; van der Kleij, Kasteleyn et al. 2019; Neves and Burgers 2022; Piera-Jiménez, Dedeu et al. 2024), or telephone contacts (Lake, Georgiou et al. 2017).

In this study, we aim to address this apparent gap in the evidence base by providing an exploratory analysis of the determinants of consultation type using a comprehensive dataset across several years. While the scope of the analysis is mainly descriptive, it would be reasonable to assume *a priori* that the choice of consultation type is partly determined by both patient- and clinic-level factors, as well as by structural factors. For example, it is broadly recognized that younger people use digital technologies to a larger extent than do older people (Odgers and Jensen 2020; Ritchie, Mathiew et al. 2023), including those for health care (Zanaboni, Nguangue et al. 2018; Cheung, Leung et al. 2019). Likewise, studies have shown that home visits are mainly conducted for patients affected by a chronic condition and with limited mobility (Clair, Sundberg et al. 2019; Sun, Parslow et al. 2022).

Obtaining an improved understanding of the factors that determine primary health care consultation type may support policy development aiming at improving the delivery of primary health care from a health systems perspective. Moreover, as different types of consultations cost differently to deliver, the findings of the analysis are relevant also from an economic perspective.

To model consultation type, we use register data on four specific primary health care consultation types – clinic visit, digital contact, home visit, telephone contact – in Sweden. The data cover all primary health care consultations in the country across the years 2019 to 2022 and includes information on both the patient and the clinic as well as on particular context factors. Sweden offers an interesting case to study these issues due to its decentralized, regionally based health care system where many factors, for example, clinic ownership and reimbursement systems, differ across clinics and regions, respectively.

The study received ethical approval from the Swedish Ethics Review Authority (EPM) on 21 September 2022 (Dnr. 2022-03964-01).

## Data and Methods

### Data

Obtaining an understanding of the factors that determine the choice of consultation type in primary health care requires access to data on several aspects of the provision of primary care. To ensure a sufficiently comprehensive dataset covering both patient and clinic as well as context factors we collected data from a range of different sources. The main data source is the Waiting-time database (sv. *Väntetidsdatabasen* (VTDB)) from the Swedish Association of Local Authorities and Regions (SALAR). The VTDB was initiated in 2019 and contains data on all primary care consultations in the country and includes all regular primary care clinics, out-of-hours clinics, and specialized clinics, such as those for maternal and newborn care.

The register contains data on the clinic, the patient, and the consultation. For the purposes of this study, data on the following indicators are used: region, clinic id (HSA-id), ownership of clinic (public or private), type of visit (new visit or re-visit), consultation type, sex, age, and home municipality of patient, diagnosis codes (ICD-10), and date of the consultation. The indicator consultation type is the outcome variable of the study and consists of four different types of primary care consultations: clinic visit; digital contact (video or chat); home visit; and phone contact. As there is no obvious reason to assume that any one of these types is inherently better than the others in any given case, these four options are treated as nominal without a natural internal ordering. The number and share of consultations by type across the study period of 2019 to 2022 are shown in Table 1.

**Table 1.** Type of consultation, 2019-2022 (N, %)

| Clinic visit | Digital contact | Home visit | Phone contact | Total |
| --- | --- | --- | --- | --- |
| **2019** |  |  |  |  |
| 10,542,028 | 55,834 | 182,684 | 922,275 | 11,702,821 |
| 90.1 | 0.5 | 1.6 | 7.9 | 100 |
| **2020** |  |  |  |  |
| 13,489,032 | 502,765 | 337,289 | 2,650,889 | 16,979,975 |
| 79.4 | 3.0 | 2.0 | 15.6 | 100 |
| **2021** |  |  |  |  |
| 15,216,949 | 774,362 | 279,630 | 3,456,420 | 19,727,361 |
| 77.1 | 3.9 | 1.4 | 17.5 | 100 |
| **2022** |  |  |  |  |
| 15,321,049 | 640,735 | 257,795 | 3,296,262 | 19,515,841 |
| 78.5 | 3.3 | 1.3 | 16.9 | 100 |

Source: SALAR VTDB 2019-2022.
Note: Two regions were missing in 2019.

In 2019, almost 12 million consultations were registered in the VTDB database. However, the actual number of consultations in that year was higher as two regions were missing in the first year of the database. In 2022, close to 20 million consultations were recorded which is a slight decrease on the previous year. Clinic visit is the dominant type of consultation across these years constituting around 80 percent of all primary care consultations. The relative fall in the share of clinic visits most likely represents a true change in the composition of consultation type across these years. Likewise, both the relative and absolute changes in the types of remote contacts by means of digital technologies and phone contacts also reflect a secular trend in the composition of the data. In particular, these types of consultations increased considerably in periods in 2020 largely as a result of the Covid-19 pandemic (Ekman, Arvidsson et al. 2021).

In addition to the VTDB, data on regional level reimbursement systems and levels have been obtained from the SALAR. Specifically, we use information on the extent to which the clinics are reimbursed for using digital contacts. This information varies across regions but not across clinics within regions.

#### Diagnoses codes

Most of the observations (consultations) contain a diagnostic code (ICD-10 code). To control for type of condition of the patient, the sample is divided into ten diagnostic code groups as described in Table 2.

**Table 2.**
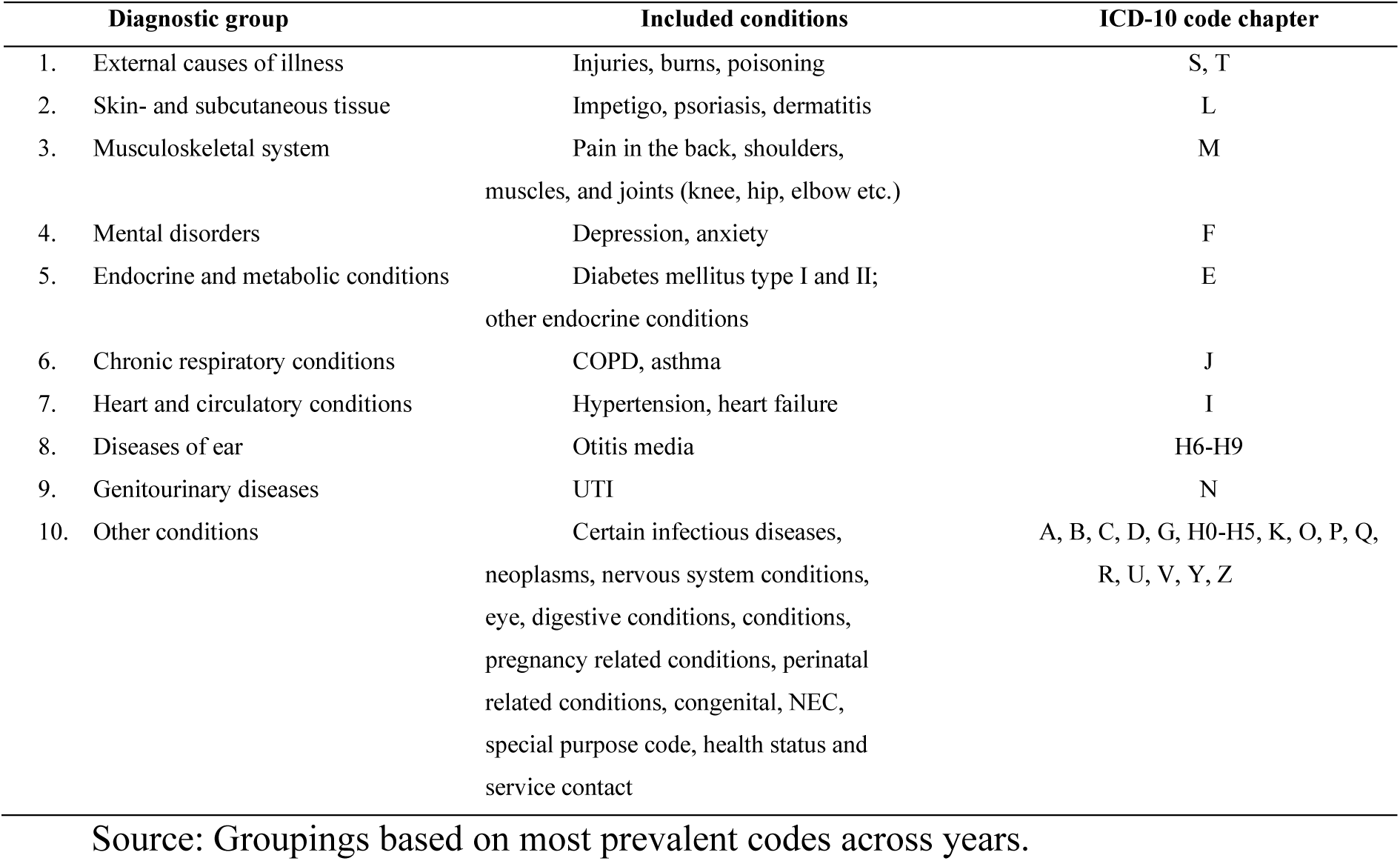
Diagnostic code groups.

| Diagnostic group |  | Included conditions | ICD-10 code chapter |
| --- | --- | --- | --- |
| 1. | External causes of illness | Injuries, burns, poisoning | S, T |
| 2. | Skin- and subcutaneous tissue | Impetigo, psoriasis, dermatitis | L |
| 3. | Musculoskeletal system | Pain in the back, shoulders, muscles, and joints (knee, hip, elbow etc.) | M |
| 4. | Mental disorders | Depression, anxiety | F |
| 5. | Endocrine and metabolic conditions | Diabetes mellitus type I and II; other endocrine conditions | E |
| 6. | Chronic respiratory conditions | COPD, asthma | J |
| 7. | Heart and circulatory conditions | Hypertension, heart failure | I |
| 8. | Diseases of ear | Otitis media | H6-H9 |
| 9. | Genitourinary diseases | UTI | N |
| 10. | Other conditions | Certain infectious diseases, neoplasms, nervous system conditions, eye, digestive conditions, conditions, pregnancy related conditions, perinatal related conditions, congenital, NEC, special purpose code, health status and service contact | A, B, C, D, G, H0-H5, K, O, P, Q, R, U, V, Y, Z |

Source: Groupings based on most prevalent codes across years.

The groupings are based on the ICD 10-chapter codes, A to Z and on the prevalence of codes in the data. To limit the number of disease groups, codes with less than around three percent prevalence have been grouped together in the *Other conditions* diagnostic group.

Descriptive statistics of the data are shown in Table 3 (see Table A.1 in Annex A for the descriptive statistics by type of consultation).

**Table 3.** Descriptive statistics, VTDB 2019-2022.

|  | 2019 | 2020 | 2021 | 2022 |
| --- | --- | --- | --- | --- |
| **Age of patient (years)** | 52<br>[25] | 52<br>[25] | 52<br>[25] | 53<br>[25] |
| **Median income of municipality adult population, SEK** | 246,207<br>[22,831] | 250,644<br>[22,743] | 264,462<br>[23,712] | 278,096<br>[24,904] |
| **Care Need Index, CNI** | 1.22<br>[0.55] | 1.31<br>[0.72] | 1.45<br>[0.80] | 1.50<br>[0.82] |
| **Adjusted Clinical Group, ACG** | 1.10<br>[0.28] | 1.07<br>[0.23] | 1.06<br>[0.21] | 1.05<br>[0.22] |
| **Number of registered patients** | 11,012<br>[4,939] | 10,105<br>[4,967] | 9,989<br>[4,924] | 9,996<br>[5,012] |
| **Number of physicians per 1,000 patients** | 0.83<br>[0.23] | 0.97<br>[0.60] | 1.02<br>[0.74] | 1.03<br>[0.76] |
| **Permanent-to-Temporary staff ratio** | 0.06<br>[0.09] | 0.06<br>[0.08] | 0.06<br>[0.08] | 0.06<br>[0.09] |
| **Share of capitation in total reimbursement, %** | 0.81<br>[0.13] | 0.86<br>[0.09] | 0.86<br>[0.08] | 0.88<br>[0.08] |
| **Reimbursement for digital contacts** | 134.26<br>[107.45] | 91.78<br>[97.23] | 84.04<br>[102.95] | 79.22<br>[101.38] |
| **Reimbursement for clinic visit** | 151.77<br>[100.79] | .<br>[.] | 79.01<br>[102.05] | .<br>[.] |
| **Sex of patient** |  |  |  |  |
| Male | 4,970,609<br>42.47 | 7,219,363<br>42.54 | 8,408,709<br>42.67 | 8,359,764<br>42.84 |
| Female | 6,732,033<br>57.53 | 9,753,394<br>57.46 | 11,295,373<br>57.33 | 11,156,077<br>57.16 |
| **New consultation** |  |  |  |  |
| Return consultation | 7,682,537<br>65.65 | 10,861,778<br>63.97 | 12,929,865<br>65.54 | 12,880,738<br>66.00 |
| New consultation | 4,020,284<br>34.35 | 6,118,197<br>36.03 | 6,797,496<br>34.46 | 6,635,103<br>34.00 |
| **Diagnosis groups** |  |  |  |  |
| External causes of illness | 297,265<br>3.87 | 488,439<br>4.10 | 511,105<br>3.69 | 499,053<br>3.59 |
| Skin and subcutaneous tissue conditions | 518,557<br>6.75 | 804,955<br>6.75 | 908,890<br>6.57 | 937,112<br>6.75 |
| Musculoskeletal system conditions | 1,086,345<br>14.14 | 1,625,070<br>13.64 | 1,799,096<br>13.00 | 1,764,357<br>12.70 |
| Mental disorders | 681,016<br>8.86 | 1,176,055<br>9.87 | 1,343,267<br>9.70 | 1,344,616<br>9.68 |
| Endocrine and metabolic conditions | 470,038<br>6.12 | 729,092<br>6.12 | 858,442<br>6.20 | 880,374<br>6.34 |
| Chronic respiratory conditions | 542,590<br>7.06 | 624,966<br>5.24 | 604,529<br>4.37 | 764,787<br>5.51 |
| Heart and circulatory conditions | 632,869<br>8.24 | 927,515<br>7.78 | 1,027,049<br>7.42 | 1,016,706<br>7.32 |
| Diseases of the ear | 269,373<br>3.51 | 373,613<br>3.13 | 425,162<br>3.07 | 484,733<br>3.49 |
| Genitourinary diseases | 224,806<br>2.93 | 360,863<br>3.03 | 398,918<br>2.88 | 413,731<br>2.98 |
| Other diseases and conditions | 2,960,293<br>38.53 | 4,807,740<br>40.34 | 5,966,124<br>43.10 | 5,785,726<br>41.65 |
| **Type of care giver** |  |  |  |  |
| Physician | 5,138,677<br>47.13 | 6,428,813<br>44.65 | 7,044,114<br>44.76 | 6,814,275<br>45.17 |
| Nurse | 2,970,925<br>27.25 | 4,420,263<br>30.70 | 4,927,112<br>31.31 | 4,637,923<br>30.74 |
| Other | 2,794,670<br>25.63 | 3,547,690<br>24.64 | 3,765,207<br>23.93 | 3,633,387<br>24.09 |
| **Public clinic** |  |  |  |  |
| Private | 2,832,555<br>24.20 | 5,155,001<br>30.36 | 7,425,911<br>37.64 | 7,563,883<br>38.76 |
| Public | 8,870,266<br>75.80 | 11,824,974<br>69.64 | 12,301,450<br>62.36 | 11,951,958<br>61.24 |

Note: Standard deviation in square brackets. Frequency and percentage share of category indicators.

In the analysis, type of consultation is the dependent variable and, depending on the type of model, a selection of these variables is used as covariates. The data contain missing observations for some variables. While no observations were removed due to missing values in the preparation of the data, missing observations are dropped by Stata to fit models.

### Methods

The aim of the analysis is to model the type of consultation in primary health care. As noted, the various alternative types of consultation – clinic visit, digital contact, home visit, telephone contact – have no natural order and can be seen as a nominal, unordered outcome. The general modelling approach for nominal outcomes is the multinomial logistic model (MNLM; Cameron and Trivedi (2022). The MNLM uses one of the alternatives as the base outcome and fits a set of binomial logistic models to estimate the probability of the outcome compared with the base alternative. We use clinic visit as the base category in the analyses.

While the data are comprehensive in their scope, the structure and nature of the data pose both opportunities and some particular issues to consider when fitting the models. First, the outcome variable – type of consultation – can be seen as the result of a choice process, the modelling of which can be done using a discrete choice model approach (Grilli and Ferrini 2022). However, such models require that the unit of analysis makes a single choice per time period (e.g., hour, day, or week). In the current case, the units (i.e., the clinics) make repeated choices per time period prohibiting the data to be choice model set (identifying cases and alternatives using cross-sectional data and identifying panels, time periods, and alternatives using panel data) before fitting the model. It would not be possible to set the time period equal to the time of the consultation, however, as that would generate an inordinate number of time periods (t = N) rendering the model impossible to fit (StataCorp 2021).

Second, the data can be seen as panel, or longitudinal, data, which would allow for particular panel data modeling approaches, such as fixed effects or random effects modeling (Frees 2004). However, also this approach is limited by the repeated outcomes per time period such that the data cannot be fully panel data set (identifying panels and time periods). We partly overcome this limitation by avoiding setting a time variable when panel data setting the data allowing us to fit a random effects panel data multinomial logistic model. And third, the clustering nature of the data would allow for a multilevel modeling approach taking into consideration the fact that consultations are nested in clinics which, in turn, are nested in regions. This would require fitting a multilevel MNLM. However, there is currently no such model available in standard econometric programs. Instead, we fit a general structural equation model (GSEM; StataCorp (2021)) which allows for fitting a two-level MNLM with random effects (Rabe-Hesketh and Skrondal 2012; Long and Freese 2014).

### The multinomial model

The general multinomial logistic model that is estimated is the following (see, e.g., Long and Freese 2014):

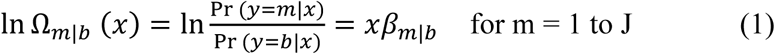

In (1), b is the ‘base category’ (or comparison group), *m* are the J alternative categories, and ***x*** is a vector of explanatory covariates. Similarly to other logit models, the multinomial logit model is interpreted as the effect of the alternative categories compared with the base category.

In the current case, the multinomial model is expressed as follows:

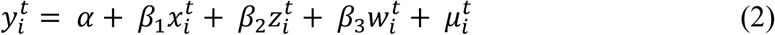

In (2), *y* is a vector that can take on the values 0, 1, 2, and 3 corresponding to the four different types of consultations (y = 0 corresponds to clinic visit and is the base category). *x* is a vector of patient level factors, including sex, age, socioeconomic status (median income of home municipality), and diagnosis group. *z* is a vector of clinic level factors, including staffing, size (as measured by the number of registered patients), and ownership of clinic. The *z* factors vary across clinics. Finally, *w* is a vector of context related factors, including reimbursement levels; these factors are constant across clinics.

### Analytical models

In the analysis, we consider these three types of structures in the data:

1. **Cross-sectional data**. Apply multinomial modeling on the cross-sectional aspect of the data using the mlogit command in Stata with appropriate options, including cluster standard errors.
2. **Panel data**. Apply panel data multinomial modeling on the panel data aspect of the data. To estimate the panel data MNLM, we first panel data set the data without a time variable and then use the xtmlogit command in Stata to fit a random effects model for valid estimates in the presence of unobserved heterogeneity at the panel level under the standard assumption of conditional independence between ***µ_i_*** and ***x_it_***.
3. **Multilevel data**. Apply multilevel multinomial modeling on the clustered aspect of the data. To estimate a two-level multilevel MNLM, we use the gsem command in Stata with a selection of the covariates.

Fitting the panel data and the multilevel MNL models are computationally demanding, including due to the curse of dimensionality (CoD; Altman and Krzywinski (2018)). To ensure convergence in the estimation the data are trimmed such that a 10 percent random sample by the clinics is obtained and, subsequently, only data from week seven is kept in each year. This week is chosen to avoid using data covering any of the Covid-19 waves that exerted an external shock to the provision of health care in Sweden (Ekman, Arvidsson et al. 2021).

## Results

This section presents the main findings of the modeling analysis. The full set of results are presented in Annex B. A set of post-estimation results, including predictions and marginal effects, are presented in Annex C.

The results of the (cross-sectional) multinomial logistic models are presented in Table 4.

**Table 4.** MNLM, Type of consultation, 2019 – 2022.

| **Clinic visit = base category** | **2019** | **2020** | **2021** | **2022** |
| --- | --- | --- | --- | --- |
| **Digital contact** |  |  |  |  |
| Sex of patient |  |  |  |  |
| Female | 0.14<br>(0.06)<br>0.016 | 0.22<br>(0.03)<br>0.000 | 0.23<br>(0.04)<br>0.000 | 0.28<br>(0.03)<br>0.000 |
| Diagnosis groups |  |  |  |  |
| Skin and subcutaneous tissue conditions | 0.41<br>(0.13)<br>0.002 | 0.07<br>(0.08)<br>0.331 | -0.08<br>(0.10)<br>0.382 | 0.12<br>(0.06)<br>0.059 |
| Musculoskeletal system conditions | 0.32<br>(0.16)<br>0.044 | 0.32<br>(0.18)<br>0.070 | 0.16<br>(0.21)<br>0.465 | 0.48<br>(0.05)<br>0.000 |
| Mental disorders | 2.10<br>(0.22)<br>0.000 | 1.96<br>(0.20)<br>0.000 | 1.91<br>(0.25)<br>0.000 | 1.97<br>(0.06)<br>0.000 |
| Endocrine and metabolic conditions | 0.64<br>(0.15)<br>0.000 | 0.97<br>(0.14)<br>0.000 | 0.87<br>(0.15)<br>0.000 | 0.94<br>(0.07)<br>0.000 |
| Chronic respiratory conditions | 0.12<br>(0.24)<br>0.625 | 1.42<br>(0.22)<br>0.000 | 0.82<br>(0.26)<br>0.002 | 0.99<br>(0.07)<br>0.000 |
| Heart and circulatory conditions | 0.59<br>(0.21)<br>0.005 | 0.79<br>(0.12)<br>0.000 | 0.70<br>(0.12)<br>0.000 | 0.84<br>(0.06)<br>0.000 |
| Diseases of the ear | -1.32<br>(0.15)<br>0.000 | -0.81<br>(0.19)<br>0.000 | -1.09<br>(0.19)<br>0.000 | -0.85<br>(0.06)<br>0.000 |
| Genitourinary diseases | 0.59<br>(0.18)<br>0.001 | 0.62<br>(0.18)<br>0.001 | 0.44<br>(0.23)<br>0.056 | 0.77<br>(0.05)<br>0.000 |
| Other diseases and conditions | 1.11<br>(0.25)<br>0.000 | 0.91<br>(0.14)<br>0.000 | 0.66<br>(0.15)<br>0.000 | 0.94<br>(0.07)<br>0.000 |
| New consultation |  |  |  |  |
| Yes | -0.17<br>(0.32)<br>0.596 | -0.24<br>(0.15)<br>0.112 | 0.16<br>(0.19)<br>0.415 | 0.08<br>(0.13)<br>0.511 |
| Age of patient (years) | -0.02<br>(0.01)<br>0.007 | -0.01<br>(0.00)<br>0.000 | -0.02<br>(0.01)<br>0.002 | -0.01<br>(0.00)<br>0.000 |
| Median income of municipality adult population, SEK | 0.00<br>(0.00) | 0.00<br>(0.00) | 0.00<br>(0.00) | 0.00<br>(0.00) |
|  | 0.000 | 0.000 | 0.000 | 0.000 |
| Intercept | -8.27<br>(0.72) | -9.64<br>(0.54) | -8.46<br>(0.60) | -7.79<br>(0.39) |
|  | 0.000 | 0.000 | 0.000 | 0.000 |
| **Home visit** |  |  |  |  |
| Sex of patient |  |  |  |  |
| Female | 0.14<br>(0.02) | 0.20<br>(0.02) | 0.19<br>(0.01) | 0.18<br>(0.01) |
|  | 0.000 | 0.000 | 0.000 | 0.000 |
| Diagnosis groups |  |  |  |  |
| Skin and subcutaneous tissue conditions | -0.17<br>(0.08) | 0.02<br>(0.06) | -0.17<br>(0.06) | -0.25<br>(0.06) |
|  | 0.035 | 0.795 | 0.002 | 0.000 |
| Musculoskeletal system conditions | -0.32<br>(0.07) | -0.34<br>(0.05) | -0.33<br>(0.05) | -0.40<br>(0.06) |
|  | 0.000 | 0.000 | 0.000 | 0.000 |
| Mental disorders | 1.51<br>(0.08) | 1.10<br>(0.07) | 1.24<br>(0.06) | 1.16<br>(0.07) |
|  | 0.000 | 0.000 | 0.000 | 0.000 |
| Endocrine and metabolic conditions | -0.27<br>(0.08) | 0.02<br>(0.06) | -0.09<br>(0.06) | -0.19<br>(0.06) |
|  | 0.000 | 0.671 | 0.119 | 0.002 |
| Chronic respiratory conditions | 0.37<br>(0.07) | 0.38<br>(0.06) | 0.41<br>(0.06) | 0.20<br>(0.07) |
|  | 0.000 | 0.000 | 0.000 | 0.004 |
| Heart and circulatory conditions | 0.13<br>(0.07) | 0.33<br>(0.05) | 0.32<br>(0.05) | 0.29<br>(0.06) |
|  | 0.064 | 0.000 | 0.000 | 0.000 |
| Diseases of the ear | -0.91<br>(0.08) | -0.72<br>(0.06) | -0.81<br>(0.08) | -0.85<br>(0.10) |
|  | 0.000 | 0.000 | 0.000 | 0.000 |
| Genitourinary diseases | -0.00<br>(0.07) | 0.02<br>(0.06) | -0.03<br>(0.05) | -0.11<br>(0.06) |
|  | 0.976 | 0.674 | 0.571 | 0.057 |
| Other diseases and conditions | 0.38<br>(0.08) | 0.38<br>(0.06) | 0.42<br>(0.06) | 0.34<br>(0.06) |
|  | 0.000 | 0.000 | 0.000 | 0.000 |
| New consultation |  |  |  |  |
| Yes | -0.61<br>(0.11) | -0.66<br>(0.07) | -0.45<br>(0.08) | -0.40<br>(0.09) |
|  | 0.000 | 0.000 | 0.000 | 0.000 |
| Age of patient<br>(years) | 0.11<br>(0.00)<br>0.000 | 0.11<br>(0.00)<br>0.000 | 0.11<br>(0.00)<br>0.000 | 0.10<br>(0.00)<br>0.000 |
| Median income of<br>municipality adult<br>population, SEK | -0.00<br>(0.00)<br>0.471 | 0.00<br>(0.00)<br>0.000 | 0.00<br>(0.00)<br>0.000 | 0.00<br>(0.00)<br>0.000 |
| Intercept | -12.27<br>(0.57)<br>0.000 | -14.46<br>(0.46)<br>0.000 | -14.20<br>(0.48)<br>0.000 | -13.45<br>(0.47)<br>0.000 |
| **Phone visit** |  |  |  |  |
| Sex of patient |  |  |  |  |
| Female | 0.14<br>(0.01)<br>0.000 | 0.16<br>(0.01)<br>0.000 | 0.13<br>(0.01)<br>0.000 | 0.13<br>(0.01)<br>0.000 |
| Diagnosis groups |  |  |  |  |
| Skin and<br>subcutaneous tissue<br>conditions | -0.50<br>(0.15)<br>0.001 | -0.20<br>(0.05)<br>0.000 | -0.12<br>(0.05)<br>0.008 | -0.03<br>(0.05)<br>0.478 |
| Musculoskeletal<br>system conditions | 0.63<br>(0.06)<br>0.000 | 0.51<br>(0.05)<br>0.000 | 0.49<br>(0.04)<br>0.000 | 0.48<br>(0.04)<br>0.000 |
| Mental disorders | 0.92<br>(0.08)<br>0.000 | 1.14<br>(0.05)<br>0.000 | 1.11<br>(0.05)<br>0.000 | 1.02<br>(0.07)<br>0.000 |
| Endocrine and<br>metabolic conditions | 0.97<br>(0.12)<br>0.000 | 1.20<br>(0.06)<br>0.000 | 1.27<br>(0.06)<br>0.000 | 1.23<br>(0.08)<br>0.000 |
| Chronic<br>respiratory conditions | 0.39<br>(0.08)<br>0.000 | 1.09<br>(0.05)<br>0.000 | 0.90<br>(0.04)<br>0.000 | 0.66<br>(0.04)<br>0.000 |
| Heart and<br>circulatory conditions | 0.50<br>(0.11)<br>0.000 | 0.81<br>(0.05)<br>0.000 | 0.83<br>(0.05)<br>0.000 | 0.87<br>(0.06)<br>0.000 |
| Diseases of the<br>ear | -0.58<br>(0.12)<br>0.000 | -0.06<br>(0.04)<br>0.127 | -0.14<br>(0.04)<br>0.001 | -0.10<br>(0.04)<br>0.022 |
| Genitourinary<br>diseases | 1.19<br>(0.07)<br>0.000 | 0.81<br>(0.04)<br>0.000 | 0.70<br>(0.04)<br>0.000 | 0.71<br>(0.04)<br>0.000 |
| Other diseases<br>and conditions | 0.81<br>(0.11)<br>0.000 | 1.07<br>(0.05)<br>0.000 | 1.04<br>(0.06)<br>0.000 | 1.12<br>(0.06)<br>0.000 |
| New consultation |  |  |  |  |
| Yes | -1.23<br>(0.33) | 0.28<br>(0.12) | 0.81<br>(0.14) | 1.04<br>(0.15) |
|  | 0.000 | 0.020 | 0.000 | 0.000 |
| Age of patient<br>(years) | 0.00<br>(0.00) | 0.00<br>(0.00) | 0.00<br>(0.00) | 0.00<br>(0.00) |
|  | 0.029 | 0.000 | 0.115 | 0.702 |
| Median income of<br>municipality adult<br>population, SEK | -0.00<br>(0.00) | 0.00<br>(0.00) | 0.00<br>(0.00) | 0.00<br>(0.00) |
|  | 0.382 | 0.212 | 0.002 | 0.000 |
| Intercept | -2.71<br>(1.18) | -3.82<br>(0.57) | -4.70<br>(0.54) | -5.14<br>(0.51) |
|  | 0.021 | 0.000 | 0.000 | 0.000 |
| N | 7,552,287 | 11,806,160 | 13,737,814 | 13,777,811 |
| AIC | 3612065 | 12935910 | 16098881 | 15374257 |
| BIC | 3612646 | 12936510 | 16099488 | 15374864 |
| Pseudo R2 | 0.098 | 0.077 | 0.080 | 0.077 |

Cluster robust standard errors
Note: SES = socioeconomic status.

Using clinic visit as the base category, digital contacts are used predominantly when the patient is younger and female. Socioeconomic status does not appear to determine the use of digital contacts in the current sample. There is a smaller probability of using digital contacts in cases involving skin and soft tissue infections compared with office visits. On the other hand, there is a larger probability of using digital contacts for cases of mental health, genitourinary, and some chronic conditions compared with clinic visits. There is some indication of a higher probability of using digital contacts among private clinics and when the clinic is reimbursed for such types of contacts by the region.

Home visits and telephone contacts are used to a larger probability for older patients and for female patients compared with clinic visits. Socioeconomic status does not affect the probability of use of these types of consultations. There is some indication that home visits are used for patients with some chronic conditions but not for patients experiencing musculoskeletal conditions. There is no indication that home visits are used for first time consultations.

The results of the panel data models are presented in Table 5.

**Table 5.** Panel data XTMNLM, Type of consultation, 2019 – 2022.

| **Clinic visit = base category** | **2019** | **2020** | **2021** | **2022** |
| --- | --- | --- | --- | --- |
| **Digital contact** |  |  |  |  |
| New consultation |  |  |  |  |
| Yes | 0.15<br>(0.35)<br>0.674 | -0.26<br>(0.25)<br>0.300 | -0.96<br>(0.17)<br>0.000 | -0.26<br>(0.12)<br>0.038 |
| Ownership of clinic |  |  |  |  |
| Public | 0.21<br>(0.41)<br>0.613 | -0.20<br>(0.27)<br>0.444 | -0.14<br>(0.16)<br>0.379 | -0.27<br>(0.14)<br>0.056 |
| Reimbursement for digital contacts |  |  |  |  |
| Yes | 1.53<br>(0.39)<br>0.000 | -0.61<br>(0.24)<br>0.011 | -0.10<br>(0.14)<br>0.476 | -0.11<br>(0.12)<br>0.362 |
| Age of patient (years) | -0.00<br>(0.00)<br>0.723 | -0.00<br>(0.00)<br>0.834 | -0.01<br>(0.00)<br>0.000 | -0.01<br>(0.00)<br>0.000 |
| Intercept | -7.94<br>(0.57)<br>0.000 | -6.03<br>(0.33)<br>0.000 | -2.82<br>(0.18)<br>0.000 | -3.00<br>(0.16)<br>0.000 |
| **Home visit** |  |  |  |  |
| New consultation |  |  |  |  |
| Yes | -0.84<br>(0.18)<br>0.000 | -0.58<br>(0.13)<br>0.000 | -0.59<br>(0.14)<br>0.000 | -0.73<br>(0.15)<br>0.000 |
| Ownership of clinic |  |  |  |  |
| Public | 0.36<br>(0.18)<br>0.051 | 0.21<br>(0.13)<br>0.115 | -0.02<br>(0.14)<br>0.904 | 0.17<br>(0.15)<br>0.266 |
| Reimbursement for digital contacts |  |  |  |  |
| Yes | -0.05<br>(0.16)<br>0.733 | -0.05<br>(0.11)<br>0.630 | 0.43<br>(0.12)<br>0.000 | 0.42<br>(0.12)<br>0.001 |
| Age of patient (years) | 0.04<br>(0.01)<br>0.000 | 0.06<br>(0.01)<br>0.000 | 0.07<br>(0.01)<br>0.000 | 0.05<br>(0.01)<br>0.000 |
| Intercept | -6.57<br>(0.52)<br>0.000 | -8.35<br>(0.47)<br>0.000 | -9.20<br>(0.53)<br>0.000 | -7.82<br>(0.45)<br>0.000 |
| **Phone visit** |  |  |  |  |
| New consultation |  |  |  |  |
| Yes | -2.10<br>(0.19)<br>0.000 | -0.15<br>(0.12)<br>0.234 | -0.12<br>(0.11)<br>0.265 | 0.22<br>(0.11)<br>0.041 |
| Ownership of clinic |  |  |  |  |
| Public | 0.13<br>(0.28)<br>0.635 | -0.93<br>(0.17)<br>0.000 | -0.75<br>(0.15)<br>0.000 | -0.65<br>(0.13)<br>0.000 |
| Reimbursement for digital contacts |  |  |  |  |
| Yes | 0.61 | -1.24 | -1.10 | -0.83 |
|  | (0.26) | (0.16) | (0.13) | (0.12) |
|  | 0.017 | 0.000 | 0.000 | 0.000 |
| Age of patient (years) | 0.01 | -0.00 | 0.00 | 0.00 |
|  | (0.00) | (0.00) | (0.00) | (0.00) |
|  | 0.000 | 0.780 | 0.000 | 0.001 |
| Intercept | -4.48 | -2.68 | -1.72 | -1.97 |
|  | (0.35) | (0.19) | (0.16) | (0.14) |
|  | 0.000 | 0.000 | 0.000 | 0.000 |
| / |  |  |  |  |
| var(u2) | 2.85 | 3.95 | 3.22 | 2.36 |
|  | (0.71) | (0.63) | (0.28) | (0.20) |
| var(u3) | 0.67 | 0.63 | 1.13 | 1.14 |
|  | (0.15) | (0.13) | (0.15) | (0.19) |
| var(u4) | 6.20 | 4.23 | 3.37 | 2.75 |
|  | (0.71) | (0.34) | (0.24) | (0.20) |
| cov(u2,u3) | -0.24 | -0.10 | 0.77 | 0.51 |
|  | (0.25) | (0.18) | (0.17) | (0.14) |
|  | 0.340 | 0.586 | 0.000 | 0.000 |
| cov(u2,u4) | 0.80 | 1.94 | 1.35 | 1.04 |
|  | (0.31) | (0.33) | (0.20) | (0.16) |
|  | 0.009 | 0.000 | 0.000 | 0.000 |
| cov(u3,u4) | -0.23 | 0.10 | 0.78 | 0.50 |
|  | (0.12) | (0.12) | (0.15) | (0.13) |
|  | 0.064 | 0.393 | 0.000 | 0.000 |
| N | 20,069 | 38,199 | 38,994 | 38,992 |
| AIC | 13081 | 24742 | 44171 | 42167 |
| BIC | 13247 | 24921 | 44351 | 42347 |

Sample trimmed to limit computational complexity.

Similar results are found employing multinomial logistic regression to the panel structure of the data. There is some evidence of correlation between the outcomes, suggesting that these models are able to adjust for some unobserved factors that also determine the outcomes and thereby the main coefficients. Notably, reimbursement for digital contacts does not appear to increase the probability of those types of contacts in this model.

The results of the multilevel MNLMs are presented in Table 6. Similar to the previous specifications, also considering the clustering structure of the data show that digital contacts are more likely for younger and female patients compared with clinic visits. The finding that there is a higher probability of digital contacts when the clinic is private remains as is the finding that reimbursements for digital contacts does not appear to affect the probability of these types of consultations.

**Table 6.** Multilevel MNLM/GSEM, type of consultation, 2019 – 2022.

| **Clinic visit = base category** | **2019** | **2020** | **2021** | **2022** |
| --- | --- | --- | --- | --- |
| **Digital contact** |  |  |  |  |
| New consultation |  |  |  |  |
| Yes | -0.55<br>(0.24)<br>0.023 | -0.01<br>(0.12)<br>0.926 | -0.19<br>(0.05)<br>0.001 | 0.18<br>(0.06)<br>0.001 |
| Ownership of clinic |  |  |  |  |
| Public | 0.53<br>(0.27)<br>0.046 | -0.70<br>(0.12)<br>0.000 | -0.59<br>(0.05)<br>0.000 | -0.41<br>(0.05)<br>0.000 |
| Reimbursement for digital contacts |  |  |  |  |
| Yes | 1.75<br>(0.32)<br>0.000 | -0.52<br>(0.12)<br>0.000 | 0.34<br>(0.05)<br>0.000 | 0.02<br>(0.05)<br>0.740 |
| Age of patient (years) | -0.00<br>(0.00)<br>0.440 | -0.00<br>(0.00)<br>0.172 | -0.01<br>(0.00)<br>0.000 | -0.01<br>(0.00)<br>0.000 |
| Intercept | -6.69<br>(0.44)<br>0.000 | -3.86<br>(0.17)<br>0.000 | -1.93<br>(0.06)<br>0.000 | -2.41<br>(0.07)<br>0.000 |
| **Home visit** |  |  |  |  |
| New consultation |  |  |  |  |
| Yes | -0.67<br>(0.15)<br>0.000 | -0.52<br>(0.10)<br>0.000 | -0.39<br>(0.10)<br>0.000 | -0.57<br>(0.12)<br>0.000 |
| Ownership of clinic |  |  |  |  |
| Public | 0.36<br>(0.16)<br>0.025 | 0.08<br>(0.10)<br>0.409 | -0.03<br>(0.09)<br>0.768 | 0.22<br>(0.10)<br>0.029 |
| Reimbursement for digital contacts |  |  |  |  |
| Yes | -0.12<br>(0.12)<br>0.317 | 0.12<br>(0.09)<br>0.163 | 0.14<br>(0.08)<br>0.079 | 0.31<br>(0.09)<br>0.000 |
| Age of patient (years) | 0.03<br>(0.00)<br>0.000 | 0.06<br>(0.00)<br>0.000 | 0.07<br>(0.00)<br>0.000 | 0.06<br>(0.00)<br>0.000 |
| Intercept | -6.11<br>(0.27)<br>0.000 | -7.83<br>(0.25)<br>0.000 | -8.21<br>(0.24)<br>0.000 | -8.22<br>(0.25)<br>0.000 |
| **Phone visit** |  |  |  |  |
| New consultation |  |  |  |  |
| Yes | -1.84<br>(0.08)<br>0.000 | -0.21<br>(0.04)<br>0.000 | 0.76<br>(0.03)<br>0.000 | 0.98<br>(0.03)<br>0.000 |
| Ownership of clinic |  |  |  |  |
| Public | 0.48<br>(0.07)<br>0.000 | -0.31<br>(0.04)<br>0.000 | -0.68<br>(0.03)<br>0.000 | -0.53<br>(0.03)<br>0.000 |
| Reimbursement for digital contacts |  |  |  |  |
| Yes | 0.13 | -1.09 | -0.71 | -0.74 |
|  | (0.05) | (0.04) | (0.03) | (0.03) |
|  | 0.011 | 0.000 | 0.000 | 0.000 |
| Age of patient (years) | 0.01 | -0.00 | 0.00 | 0.00 |
|  | (0.00) | (0.00) | (0.00) | (0.00) |
|  | 0.000 | 0.578 | 0.000 | 0.000 |
| Intercept | -2.87 | -1.64 | -1.23 | -1.55 |
|  | (0.09) | (0.06) | (0.04) | (0.04) |
|  | 0.000 | 0.000 | 0.000 | 0.000 |
| N | 20,055 | 38,206 | 39,001 | 38,723 |
| AIC | 15732 | 28177 | 54015 | 49548 |
| BIC | 15850 | 28305 | 54143 | 49676 |
| Sample trimmed to avoid computational complexity. |  |  |  |  |

There is a strong probability of home visits when the patient is older, but a smaller probability of such consultations when the clinic is public. Similar to digital contacts, the probability of phone consultations increases when the patient is female, but contrary to the former types of consultations the probability increases by age compared with the base category of clinic visits.

### Additional findings

In addition to the main findings, a set of extended analyses were made (see Annex B for details). First, the cross-sectional MNLM was refitted by type of ownership (public or private). We found, among other things, that there is a difference between public and private clinics in the probability of using digital contacts where public clinics have a lower probability of using that type of consultation compared with clinic visits while private clinics have a higher probability. These findings are in line with the main findings reported above.

We also find that while both types of ownerships appear to lead to an increased probability of using digital contacts operating under a reimbursement regime that includes payments for digital consultations, the effect is larger for private clinics. However, while there is a positive effect on the probability of using digital contacts when these are included in the reimbursement scheme, there is no evidence that this effect is either strong or consistent across reimbursement levels as shown in Figure C.3 Predicted margins, digital reimbursement, 2019-2022.

## Discussion

The results of the modelling of consultation type in the current context suggest that the type of primary health care consultation is determined by both patient and clinic factors as well as context characteristics. They also show that different types of consultations are determined by different factors and sometimes by the same factors having the opposite effect. While the use of digital health technologies, also in the area of primary health care have been the subject of investigation over the past decade or so (see, e.g., Torre-Diez, López-Coronado et al. 2015; Zanaboni, Nguangue et al. 2018), the use of other types of primary health care consultations, such as home visits (Marcinowicz, Chlabicz et al. 2007; Sun, Parslow et al. 2022) and telephone contacts (Lake, Georgiou et al. 2017) and how they relate to the use of clinic visits has not been the object of investigation to the same extent. Importantly, by analysing the full spectrum of consultations, the results of this study may provide some further evidence into the application of primary health care in an advanced setting. The finding that directed reimbursements for digital contacts only has a small and largely incoherent effect is noteworthy.

The heterogeneity across the models and factors indicates the complex nature of these outcomes. As noted earlier, the choice of consultation type is likely to be the result of a combination of several factors, both on the patient and the provider side. However, the mixed findings may also point at a lack of a clear strategy for how to make the most effective use of the available alternatives for treating patients in primary health care. The absence of such a strategy may have implications on the overall costs of care and on the ability to make primary health care more patient centered.

Previous research has shown that the cost of delivering a unit of care varies by type of consultation. For example, Ekman (2017) found that the average unit cost of a clinic visit was estimated at SEK 1,500 compared with SEK 800 for a digital contact in the Swedish context.

Similar differences have been found also in other countries and settings (Kouskoukis and Botsaris 2017). This suggests that there is scope for primary care providers to direct certain types of patients to different types of consultations to ensure efficient use of resources.

An additional issue of relevance for further study is the question of whether the introduction of digital contacts has an effect on the overall supply of primary care. In other words, does the application of digital health technologies improve the providers’ productivity such that more care is produced for a given allocation of financial and other resources (Young, Nesbitt et al. 2017). Relatedly, digital technologies for health care are likely to become increasingly more enabled by artificial intelligence (AI; such as machine learning, large language models (LLMs), and automation of decisions). While primary health care still lags behind other medical fields in this regard, such as medical imaging, oncology, and cardiology, AI technologies are likely to become more prevalent also in primary health care.

The study is affected by a series of limitations. First, given the idiosyncratic nature of health care services, the study setting of Sweden may limit generalizability somewhat. Focusing on the broader findings is therefore advisable. Second, interpretability of the estimates needs to be made with care. For example, the coefficient estimates of the panel data MNL models are defined as the natural logarithm of the relative risk (i.e., a double ratio) relative to the base category. It is preferable to focus on the signs of these estimates when interpreting results. We also present the predicted margins for a proposed policy variable (reimbursement) for enhanced interpretability. And third, to arrive at a more general case we fit the panel data MNL under the assumption of an independent covariance structure and an unstructured matrix.

## Conclusions

The types of primary health care consultations are determined by both patient, clinic, as well as context level factors. While policy makers are unable to directly influence the patient level factors, they can introduce incentives and regulations that may benefit both patients and the effectiveness of the overall primary care system by maximizing the various types of consultations across the varying needs of patients. To further enhance the use of digital consultations, incentives may need to be strengthened. While the clinic visit is likely to continue being the dominant form of consultation in primary health care, understanding what factors drive the use of other types of consultations may support the development of strategies both at the clinic and the overall system level.

## Data Availability

All data produced in the present study are available upon reasonable request to the authors.

## Annex A. Descriptive statistics

|  | Type of consultation, 2019 |  |  |  | Total |
| --- | --- | --- | --- | --- | --- |
|  | Clinic visit | Digital contact | Home visit | Phone contact |  |
| Age of patient (years) | 51<br>[25] | 45<br>[25] | 71<br>[30] | 56<br>[23] | 52<br>[25] |
| Median income of municipality adult population, SEK | 246,399<br>[23,082] | 257,663<br>[24,937] | 248,428<br>[25,570] | 242,909<br>[18,424] | 246,207<br>[22,831] |
| Care Need Index, CNI | 1.20<br>[0.54] | 1.09<br>[0.47] | 1.21<br>[0.51] | 1.43<br>[0.69] | 1.22<br>[0.55] |
| Adjusted Clinical Group, ACG | 1.11<br>[0.29] | 1.01<br>[0.14] | 1.09<br>[0.25] | 1.02<br>[0.11] | 1.10<br>[0.28] |
| Number of registered patients | 11,035<br>[4,964] | 10,150<br>[4,853] | 11,086<br>[5,126] | 10,787<br>[4,590] | 11,012<br>[4,939] |
| Number of physicians per 1,000 patients | 0.83<br>[0.23] | 0.82<br>[0.19] | 0.81<br>[0.23] | 0.85<br>[0.21] | 0.83<br>[0.23] |
| Permanent-to-Temporary staff ratio | 0.06<br>[0.09] | 0.02<br>[0.05] | 0.06<br>[0.09] | 0.07<br>[0.11] | 0.06<br>[0.09] |
| Share of capitation in total reimbursement, % | 0.80<br>[0.13] | 0.87<br>[0.10] | 0.80<br>[0.13] | 0.81<br>[0.09] | 0.81<br>[0.13] |
| Reimbursement for digital contacts | 134.52<br>[107.10] | 183.91<br>[66.10] | 134.96<br>[107.70] | 128.08<br>[112.41] | 134.26<br>[107.45] |
| Reimbursement for clinic visit | 152.62<br>[98.36] | 184.81<br>[65.79] | 154.25<br>[92.99] | 139.57<br>[126.91] | 151.77<br>[100.79] |
| Sex of patient |  |  |  |  |  |
| Male | 4,504,797<br>42.73 | 20,781<br>37.22 | 75,377<br>41.26 | 369,654<br>40.08 | 4,970,609<br>42.47 |
| Female | 6,037,062<br>57.27 | 35,053<br>62.78 | 107,307<br>58.74 | 552,611<br>59.92 | 6,732,033<br>57.53 |
| New consultation |  |  |  |  |  |
| Return consultation | 6,671,797<br>63.29 | 37,809<br>67.72 | 152,906<br>83.70 | 820,025<br>88.91 | 7,682,537<br>65.65 |
| New consultation | 3,870,231<br>36.71 | 18,025<br>32.28 | 29,778<br>16.30 | 102,250<br>11.09 | 4,020,284<br>34.35 |
| Diagnosis groups |  |  |  |  |  |
| External causes of illness | 289,433<br>3.99 | 593<br>1.51 | 2,352<br>2.46 | 4,887<br>1.63 | 297,265<br>3.87 |
| Skin and subcutaneous tissue conditions | 504,578<br>6.96 | 1,545<br>3.93 | 6,022<br>6.29 | 6,412<br>2.14 | 518,557<br>6.75 |
| Musculoskeletal system conditions | 1,033,969<br>14.26 | 2,436<br>6.19 | 7,265<br>7.59 | 42,675<br>14.25 | 1,086,345<br>14.14 |
| Mental disorders | 619,544<br>8.55 | 12,412<br>31.53 | 11,245<br>11.75 | 37,815<br>12.62 | 681,016<br>8.86 |
| Endocrine and metabolic conditions | 431,756<br>5.96 | 1,193<br>3.03 | 5,200<br>5.43 | 31,889<br>10.65 | 470,038<br>6.12 |
| Chronic respiratory conditions | 524,904<br>7.24 | 1,451<br>3.69 | 4,132<br>4.32 | 12,103<br>4.04 | 542,590<br>7.06 |
| Heart and circulatory conditions | 590,982<br>8.15 | 1,417<br>3.60 | 15,446<br>16.14 | 25,024<br>8.35 | 632,869<br>8.24 |
| Diseases of the ear | 265,627<br>3.66 | 162<br>0.41 | 1,082<br>1.13 | 2,502<br>0.84 | 269,373<br>3.51 |
| Genitourinary diseases | 209,416<br>2.89 | 664<br>1.69 | 2,663<br>2.78 | 12,063<br>4.03 | 224,806<br>2.93 |
| Other diseases and conditions | 2,778,349<br>38.33 | 17,487<br>44.43 | 40,280<br>42.10 | 124,177<br>41.45 | 2,960,293<br>38.53 |
| Type of care giver |  |  |  |  |  |
| Physician | 4,604,778<br>46.30 | 30,077<br>54.30 | 68,685<br>38.26 | 435,137<br>60.13 | 5,138,677<br>47.13 |
| Nurse | 2,716,257<br>27.31 | 12,051<br>21.76 | 32,030<br>17.84 | 210,587<br>29.10 | 2,970,925<br>27.25 |
| Other | 2,624,614<br>26.39 | 13,260<br>23.94 | 78,825<br>43.90 | 77,971<br>10.77 | 2,794,670<br>25.63 |
| Public clinic |  |  |  |  |  |
| Private | 2,595,367<br>24.62 | 17,946<br>32.14 | 37,728<br>20.65 | 181,514<br>19.68 | 2,832,555<br>24.20 |
| Public | 7,946,661<br>75.38 | 37,888<br>67.86 | 144,956<br>79.35 | 740,761<br>80.32 | 8,870,266<br>75.80 |
| Total number of consultations | 10,542,028 | 55,834 | 182,684 | 922,275 | 11,702,821 |

|  | Type of consultation, 2020 |  |  |  | Total |
| --- | --- | --- | --- | --- | --- |
|  | Clinic visit | Digital contact | Home visit | Phone contact |  |
| Age of patient (years) | 51<br>[25] | 45<br>[22] | 76<br>[23] | 53<br>[24] | 52<br>[25] |
| Median income of municipality adult population, SEK | 249,845<br>[22,638] | 265,842<br>[27,678] | 255,850<br>[27,558] | 251,141<br>[20,317] | 250,644<br>[22,743] |
| Care Need Index, CNI | 1.28<br>[0.67] | 1.22<br>[0.62] | 1.34<br>[0.73] | 1.52<br>[0.89] | 1.31<br>[0.72] |
| Adjusted Clinical Group, ACG | 1.08<br>[0.25] | 1.01<br>[0.12] | 1.10<br>[0.26] | 1.03<br>[0.15] | 1.07<br>[0.23] |
| Number of registered patients | 10,152<br>[4,992] | 10,971<br>[5,932] | 10,531<br>[5,001] | 9,648<br>[4,584] | 10,105<br>[4,967] |
| Number of physicians per 1,000 patients | 0.96<br>[0.58] | 0.89<br>[0.57] | 0.90<br>[0.45] | 1.01<br>[0.71] | 0.97<br>[0.60] |
| Permanent-to-Temporary staff ratio | 0.06<br>[0.08] | 0.03<br>[0.07] | 0.05<br>[0.08] | 0.06<br>[0.08] | 0.06<br>[0.08] |
| Share of capitation in total reimbursement, % | 0.86<br>[0.09] | 0.83<br>[0.11] | 0.84<br>[0.09] | 0.87<br>[0.07] | 0.86<br>[0.09] |
| Reimbursement for digital contacts | 96.39<br>[97.06] | 114.18<br>[90.04] | 103.84<br>[93.70] | 62.50<br>[94.31] | 91.78<br>[97.23] |
| Reimbursement for clinic visit | .<br>[.] | .<br>[.] | .<br>[.] | .<br>[.] | .<br>[.] |
| Sex of patient |  |  |  |  |  |
| Male | 5,847,182<br>43.35 | 182,146<br>36.29 | 136,198<br>40.39 | 1,053,837<br>39.82 | 7,219,363<br>42.54 |
| Female | 7,640,123<br>56.65 | 319,808<br>63.71 | 201,017<br>59.61 | 1,592,446<br>60.18 | 9,753,394<br>57.46 |
| New consultation |  |  |  |  |  |
| Return consultation | 8,705,887<br>64.54 | 343,049<br>68.23 | 282,343<br>83.71 | 1,530,499<br>57.74 | 10,861,778<br>63.97 |
| New consultation | 4,783,145<br>35.46 | 159,716<br>31.77 | 54,946<br>16.29 | 1,120,390<br>42.26 | 6,118,197<br>36.03 |
| Diagnosis groups |  |  |  |  |  |
| External causes of illness | 449,194<br>4.54 | 6,645<br>1.65 | 6,052<br>2.92 | 26,548<br>1.88 | 488,439<br>4.10 |
| Skin and subcutaneous tissue conditions | 739,553<br>7.47 | 12,551<br>3.12 | 16,809<br>8.10 | 36,042<br>2.55 | 804,955<br>6.75 |
| Musculoskeletal system conditions | 1,440,110<br>14.55 | 27,686<br>6.88 | 14,755<br>7.11 | 142,519<br>10.10 | 1,625,070<br>13.64 |
| Mental disorders | 882,772<br>8.92 | 118,224<br>29.37 | 17,126<br>8.25 | 157,933<br>11.19 | 1,176,055<br>9.87 |
| Endocrine and metabolic conditions | 585,624<br>5.92 | 20,164<br>5.01 | 13,639<br>6.57 | 109,665<br>7.77 | 729,092<br>6.12 |
| Chronic respiratory conditions | 492,984<br>4.98 | 35,589<br>8.84 | 6,288<br>3.03 | 90,105<br>6.39 | 624,966<br>5.24 |
| Heart and circulatory conditions | 768,134<br>7.76 | 21,164<br>5.26 | 36,919<br>17.78 | 101,298<br>7.18 | 927,515<br>7.78 |
| Diseases of the ear | 348,038<br>3.52 | 2,488<br>0.62 | 2,925<br>1.41 | 20,162<br>1.43 | 373,613<br>3.13 |
| Genitourinary diseases | 303,518<br>3.07 | 7,853<br>1.95 | 6,537<br>3.15 | 42,955<br>3.04 | 360,863<br>3.03 |
| Other diseases and conditions | 3,887,315<br>39.28 | 150,132<br>37.30 | 86,558<br>41.69 | 683,735<br>48.46 | 4,807,740<br>40.34 |
| Type of care giver |  |  |  |  |  |
| Physician | 5,149,707<br>45.61 | 292,258<br>61.24 | 84,066<br>30.08 | 902,782<br>38.43 | 6,428,813<br>44.65 |
| Nurse | 3,172,209<br>28.09 | 71,954<br>15.08 | 72,797<br>26.05 | 1,103,303<br>46.97 | 4,420,263<br>30.70 |
| Other | 2,969,287<br>26.30 | 113,040<br>23.69 | 122,586<br>43.87 | 342,777<br>14.59 | 3,547,690<br>24.64 |
| Public clinic |  |  |  |  |  |
| Private | 3,688,800<br>27.35 | 215,929<br>42.95 | 79,171<br>23.47 | 1,171,101<br>44.18 | 5,155,001<br>30.36 |
| Public | 9,800,232<br>72.65 | 286,836<br>57.05 | 258,118<br>76.53 | 1,479,788<br>55.82 | 11,824,974<br>69.64 |
| Total number of consultations | 13,489,032 | 502,765 | 337,289 | 2,650,889 | 16,979,975 |

|  | Type of consultation, 2021 |  |  |  | Total |
| --- | --- | --- | --- | --- | --- |
|  | Clinic visit | Digital contact | Home visit | Phone contact |  |
| Age of patient (years) | 52<br>[25] | 43<br>[23] | 75<br>[25] | 52<br>[24] | 52<br>[25] |
| Median income of municipality adult population, SEK | 263,355<br>[23,706] | 277,078<br>[27,381] | 268,962<br>[28,896] | 266,122<br>[21,305] | 264,462<br>[23,712] |
| Care Need Index, CNI | 1.39<br>[0.74] | 1.43<br>[0.79] | 1.33<br>[0.74] | 1.71<br>[0.99] | 1.45<br>[0.80] |
| Adjusted Clinical Group, ACG | 1.07<br>[0.23] | 1.01<br>[0.13] | 1.07<br>[0.21] | 1.03<br>[0.15] | 1.06<br>[0.21] |
| Number of registered patients | 10,023<br>[4,926] | 10,865<br>[5,634] | 10,209<br>[5,109] | 9,625<br>[4,694] | 9,989<br>[4,924] |
| Number of physicians per 1,000 patients | 1.03<br>[0.73] | 0.89<br>[0.55] | 0.97<br>[0.68] | 1.03<br>[0.80] | 1.02<br>[0.74] |
| Permanent-to-Temporary staff ratio | 0.06<br>[0.08] | 0.04<br>[0.06] | 0.05<br>[0.09] | 0.05<br>[0.08] | 0.06<br>[0.08] |
| Share of capitation in total reimbursement, % | 0.87<br>[0.08] | 0.85<br>[0.09] | 0.86<br>[0.08] | 0.85<br>[0.07] | 0.86<br>[0.08] |
| Reimbursement for digital contacts | 90.64<br>[103.70] | 81.36<br>[87.99] | 99.50<br>[98.58] | 54.34<br>[97.64] | 84.04<br>[102.95] |
| Reimbursement for clinic visit | 84.23<br>[102.92] | 80.52<br>[87.81] | 97.61<br>[98.55] | 54.19<br>[97.57] | 79.01<br>[102.05] |
| Sex of patient |  |  |  |  |  |
| Male | 6,627,518<br>43.59 | 277,811<br>35.95 | 111,412<br>39.86 | 1,391,968<br>40.39 | 8,408,709<br>42.67 |
| Female | 8,577,907<br>56.41 | 494,947<br>64.05 | 168,095<br>60.14 | 2,054,424<br>59.61 | 11,295,373<br>57.33 |
| New consultation |  |  |  |  |  |
| Return consultation | 10,524,357<br>69.16 | 499,473<br>64.50 | 230,970<br>82.60 | 1,675,065<br>48.46 | 12,929,865<br>65.54 |
| New consultation | 4,692,592<br>30.84 | 274,889<br>35.50 | 48,660<br>17.40 | 1,781,355<br>51.54 | 6,797,496<br>34.46 |
| Diagnosis groups |  |  |  |  |  |
| External causes of illness | 458,653<br>4.09 | 12,909<br>2.03 | 4,867<br>2.66 | 34,676<br>1.91 | 511,105<br>3.69 |
| Skin and subcutaneous tissue conditions | 824,887<br>7.36 | 20,781<br>3.27 | 12,092<br>6.61 | 51,130<br>2.81 | 908,890<br>6.57 |
| Musculoskeletal system conditions | 1,567,128<br>13.98 | 42,907<br>6.76 | 13,105<br>7.16 | 175,956<br>9.68 | 1,799,096<br>13.00 |
| Mental disorders | 951,921<br>8.49 | 194,478<br>30.63 | 17,138<br>9.37 | 179,730<br>9.89 | 1,343,267<br>9.70 |
| Endocrine and metabolic conditions | 675,364<br>6.03 | 31,865<br>5.02 | 10,564<br>5.77 | 140,649<br>7.74 | 858,442<br>6.20 |
| Chronic respiratory conditions | 471,662<br>4.21 | 36,773<br>5.79 | 5,162<br>2.82 | 90,932<br>5.00 | 604,529<br>4.37 |
| Heart and circulatory conditions | 843,071<br>7.52 | 31,919<br>5.03 | 29,473<br>16.11 | 122,586<br>6.74 | 1,027,049<br>7.42 |
| Diseases of the ear | 391,905<br>3.50 | 4,062<br>0.64 | 2,428<br>1.33 | 26,767<br>1.47 | 425,162<br>3.07 |
| Genitourinary diseases | 328,972<br>2.94 | 12,647<br>1.99 | 5,474<br>2.99 | 51,825<br>2.85 | 398,918<br>2.88 |
| Other diseases and conditions | 4,693,256<br>41.88 | 246,581<br>38.84 | 82,653<br>45.18 | 943,634<br>51.91 | 5,966,124<br>43.10 |
| Type of care giver |  |  |  |  |  |
| Physician | 5,566,240<br>47.12 | 339,510<br>51.59 | 82,861<br>34.17 | 1,055,503<br>34.92 | 7,044,114<br>44.76 |
| Nurse | 3,259,326<br>27.59 | 142,151<br>21.60 | 45,389<br>18.72 | 1,480,246<br>48.97 | 4,927,112<br>31.31 |
| Other | 2,987,647<br>25.29 | 176,371<br>26.80 | 114,215<br>47.11 | 486,974<br>16.11 | 3,765,207<br>23.93 |
| Public clinic |  |  |  |  |  |
| Private | 5,130,745<br>33.72 | 356,662<br>46.06 | 77,357<br>27.66 | 1,861,147<br>53.85 | 7,425,911<br>37.64 |
| Public | 10,086,204<br>66.28 | 417,700<br>53.94 | 202,273<br>72.34 | 1,595,273<br>46.15 | 12,301,450<br>62.36 |
| Total number of consultations | 15,216,949 | 774,362 | 279,630 | 3,456,420 | 19,727,361 |

|  | Type of consultation, 2022 |  |  |  | Total |
| --- | --- | --- | --- | --- | --- |
|  | Clinic visit | Digital contact | Home visit | Phone contact |  |
| Age of patient (years) | 53<br>[25] | 47<br>[21] | 74<br>[26] | 52<br>[24] | 53<br>[25] |
| Median income of municipality adult population, SEK | 277,143<br>[24,959] | 286,815<br>[27,922] | 280,991<br>[28,771] | 280,581<br>[23,120] | 278,096<br>[24,904] |
| Care Need Index, CNI | 1.44<br>[0.77] | 1.47<br>[0.81] | 1.38<br>[0.78] | 1.77<br>[0.97] | 1.50<br>[0.82] |
| Adjusted Clinical Group, ACG | 1.06<br>[0.24] | 1.01<br>[0.14] | 1.06<br>[0.22] | 1.02<br>[0.13] | 1.05<br>[0.22] |
| Number of registered patients | 10,011<br>[5,004] | 10,804<br>[5,677] | 10,087<br>[5,127] | 9,759<br>[4,883] | 9,996<br>[5,012] |
| Number of physicians per 1,000 patients | 1.03<br>[0.74] | 0.96<br>[0.68] | 0.97<br>[0.63] | 1.05<br>[0.84] | 1.03<br>[0.76] |
| Permanent-to-Temporary staff ratio | 0.06<br>[0.09] | 0.04<br>[0.07] | 0.06<br>[0.09] | 0.06<br>[0.08] | 0.06<br>[0.09] |
| Share of capitation in total reimbursement, % | 0.88<br>[0.08] | 0.88<br>[0.09] | 0.88<br>[0.08] | 0.89<br>[0.07] | 0.88<br>[0.08] |
| Reimbursement for digital contacts | 84.62<br>[101.41] | 75.04<br>[89.17] | 100.86<br>[102.10] | 53.25<br>[99.19] | 79.22<br>[101.38] |
| Reimbursement for clinic visit | .<br>[.] | .<br>[.] | .<br>[.] | .<br>[.] | .<br>[.] |
| Sex of patient |  |  |  |  |  |
| Male | 6,700,166<br>43.73 | 225,211<br>35.15 | 104,423<br>40.51 | 1,329,964<br>40.35 | 8,359,764<br>42.84 |
| Female | 8,620,883<br>56.27 | 415,524<br>64.85 | 153,372<br>59.49 | 1,966,298<br>59.65 | 11,156,077<br>57.16 |
| New consultation |  |  |  |  |  |
| Return consultation | 10,780,521<br>70.36 | 431,069<br>67.28 | 215,238<br>83.49 | 1,453,910<br>44.11 | 12,880,738<br>66.00 |
| New consultation | 4,540,528<br>29.64 | 209,666<br>32.72 | 42,557<br>16.51 | 1,842,352<br>55.89 | 6,635,103<br>34.00 |
| Diagnosis groups |  |  |  |  |  |
| External causes of illness | 453,286<br>3.98 | 7,569<br>1.49 | 4,770<br>2.85 | 33,428<br>1.84 | 499,053<br>3.59 |
| Skin and subcutaneous tissue conditions | 854,230<br>7.50 | 16,345<br>3.21 | 11,394<br>6.82 | 55,143<br>3.03 | 937,112<br>6.75 |
| Musculoskeletal system conditions | 1,548,036<br>13.58 | 39,360<br>7.73 | 12,471<br>7.46 | 164,490<br>9.04 | 1,764,357<br>12.70 |
| Mental disorders | 1,020,593<br>8.96 | 142,416<br>27.99 | 16,282<br>9.74 | 165,325<br>9.09 | 1,344,616<br>9.68 |
| Endocrine and metabolic conditions | 713,801<br>6.26 | 26,511<br>5.21 | 9,883<br>5.91 | 130,179<br>7.16 | 880,374<br>6.34 |
| Chronic respiratory conditions | 627,471<br>5.51 | 33,049<br>6.49 | 5,373<br>3.22 | 98,894<br>5.44 | 764,787<br>5.51 |
| Heart and circulatory conditions | 844,625<br>7.41 | 27,492<br>5.40 | 27,161<br>16.25 | 117,428<br>6.46 | 1,016,706<br>7.32 |
| Diseases of the ear | 446,568<br>3.92 | 3,525<br>0.69 | 2,445<br>1.46 | 32,195<br>1.77 | 484,733<br>3.49 |
| Genitourinary diseases | 343,387<br>3.01 | 12,060<br>2.37 | 5,142<br>3.08 | 53,142<br>2.92 | 413,731<br>2.98 |
| Other diseases and conditions | 4,544,353<br>39.88 | 200,561<br>39.41 | 72,186<br>43.20 | 968,626<br>53.25 | 5,785,726<br>41.65 |
| Type of care giver |  |  |  |  |  |
| Physician | 5,507,608<br>47.86 | 306,261<br>58.70 | 75,574<br>33.92 | 924,832<br>32.64 | 6,814,275<br>45.17 |
| Nurse | 3,065,215<br>26.64 | 95,038<br>18.21 | 35,585<br>15.97 | 1,442,085<br>50.90 | 4,637,923<br>30.74 |
| Other | 2,935,084<br>25.50 | 120,474<br>23.09 | 111,641<br>50.11 | 466,188<br>16.46 | 3,633,387<br>24.09 |
| Public clinic |  |  |  |  |  |
| Private | 5,470,398<br>35.71 | 263,779<br>41.17 | 67,554<br>26.20 | 1,762,152<br>53.46 | 7,563,883<br>38.76 |
| Public | 9,850,651<br>64.29 | 376,956<br>58.83 | 190,241<br>73.80 | 1,534,110<br>46.54 | 11,951,958<br>61.24 |
| Total number of consultations | 15,321,049 | 640,735 | 257,795 | 3,296,262 | 19,515,841 |

## Annex B. Full estimation results

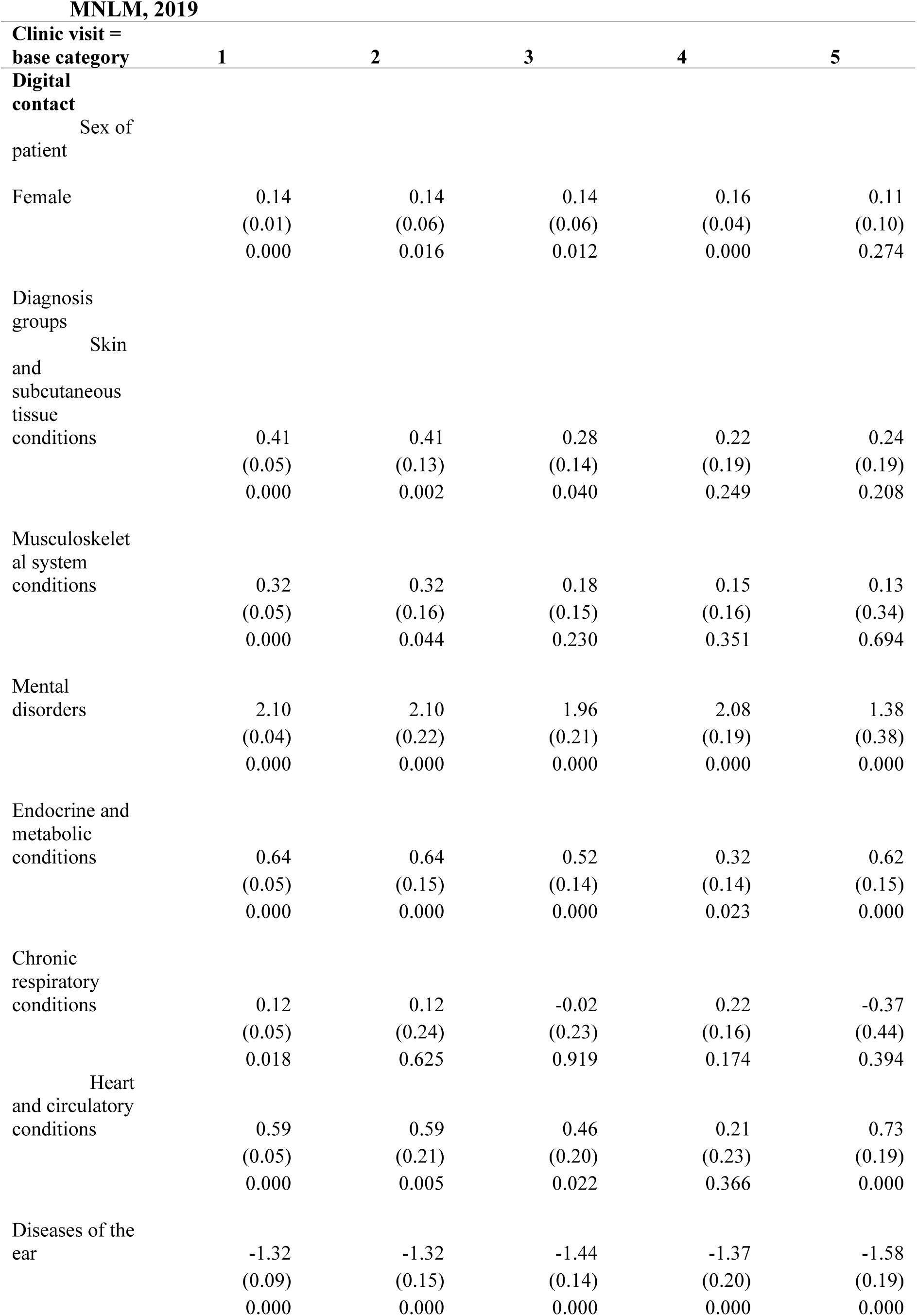
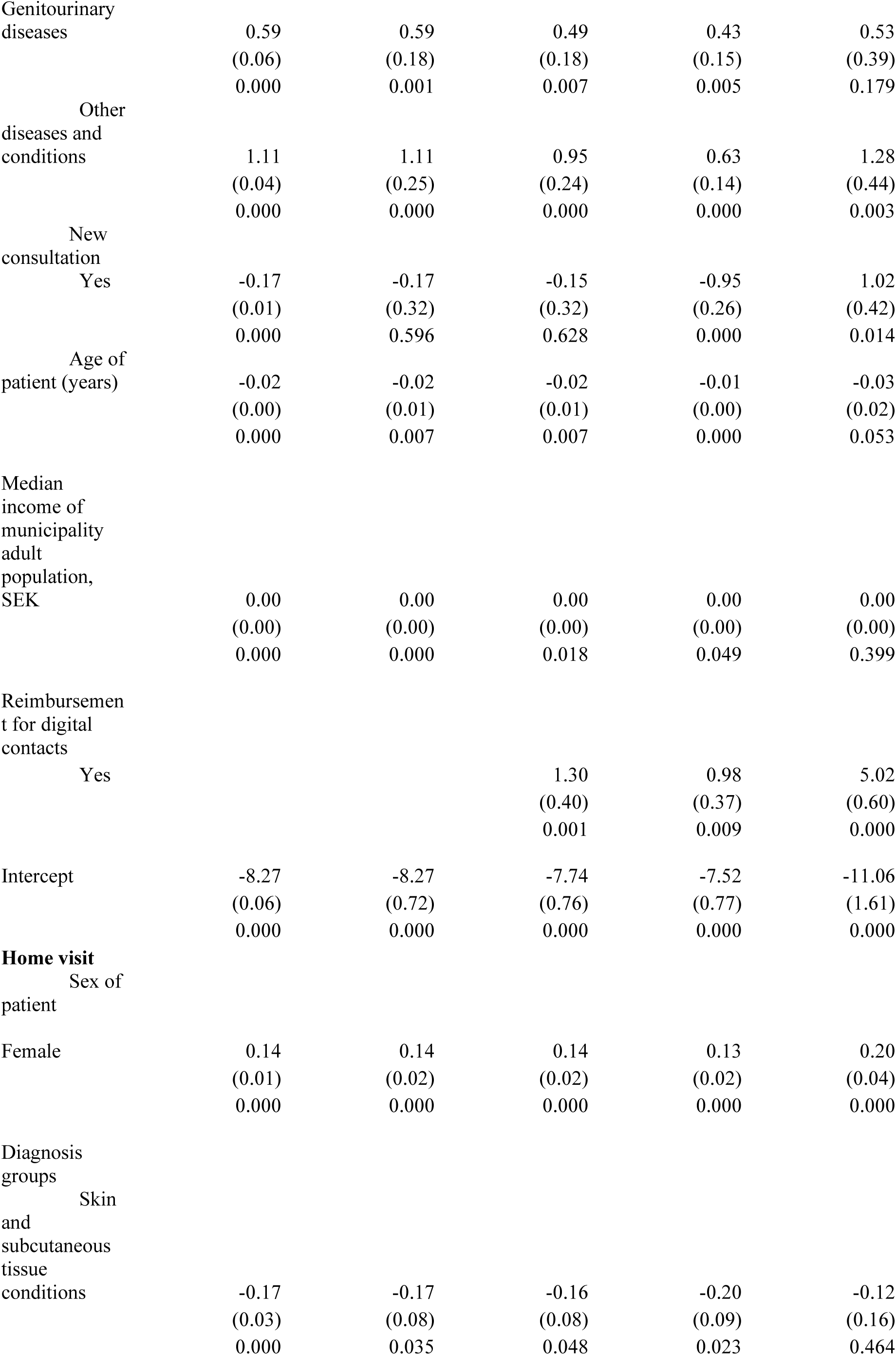
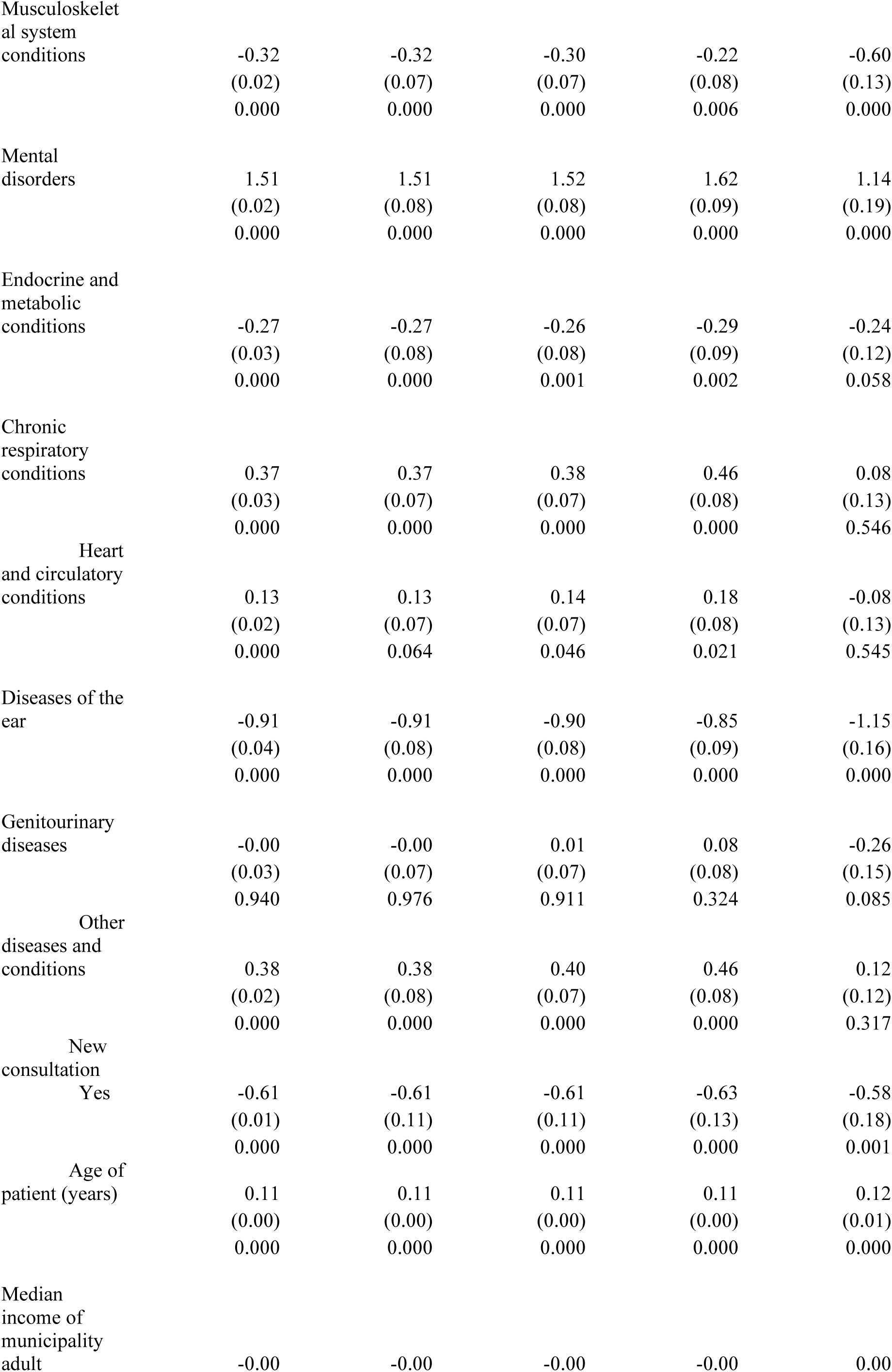
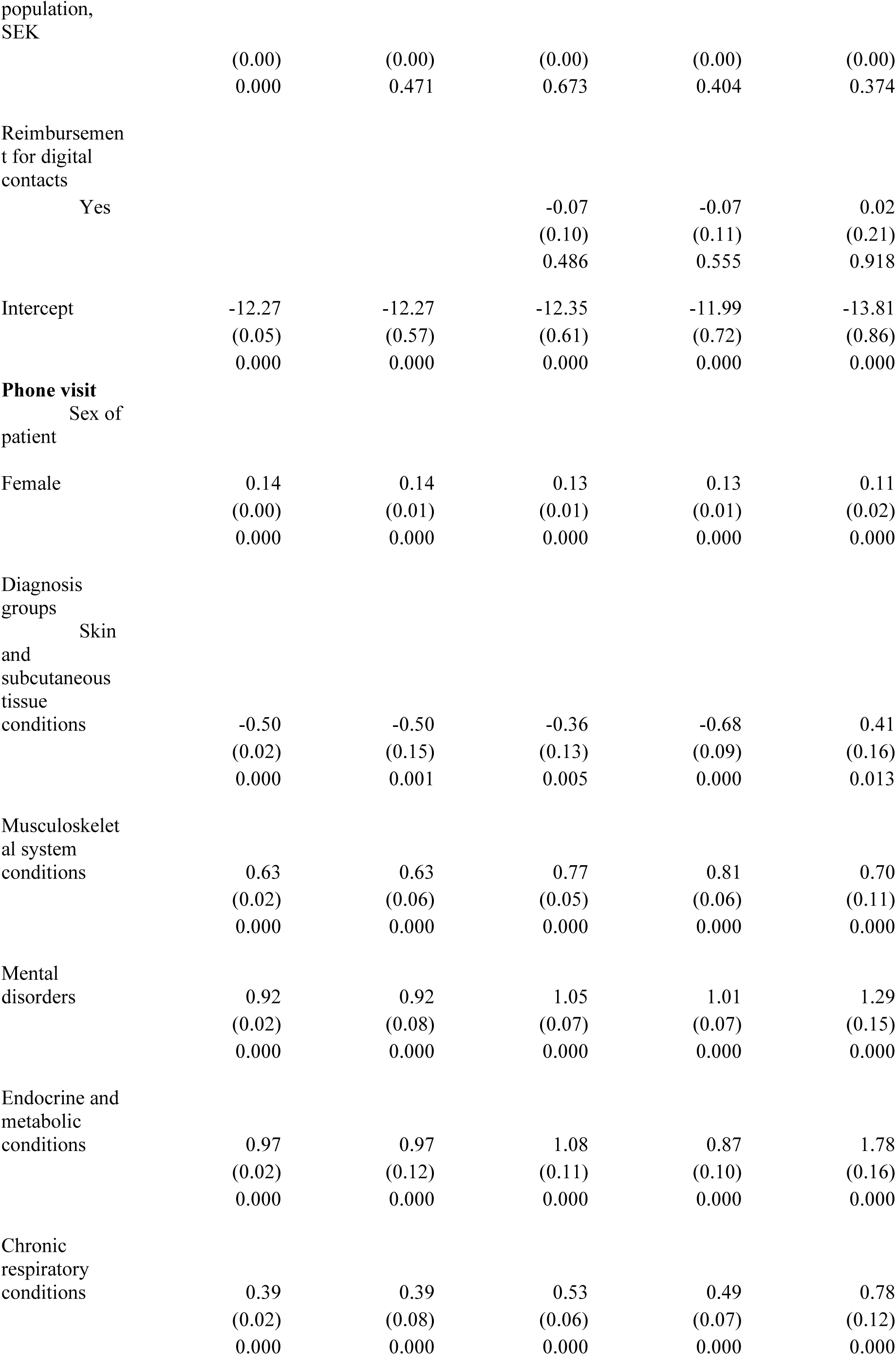
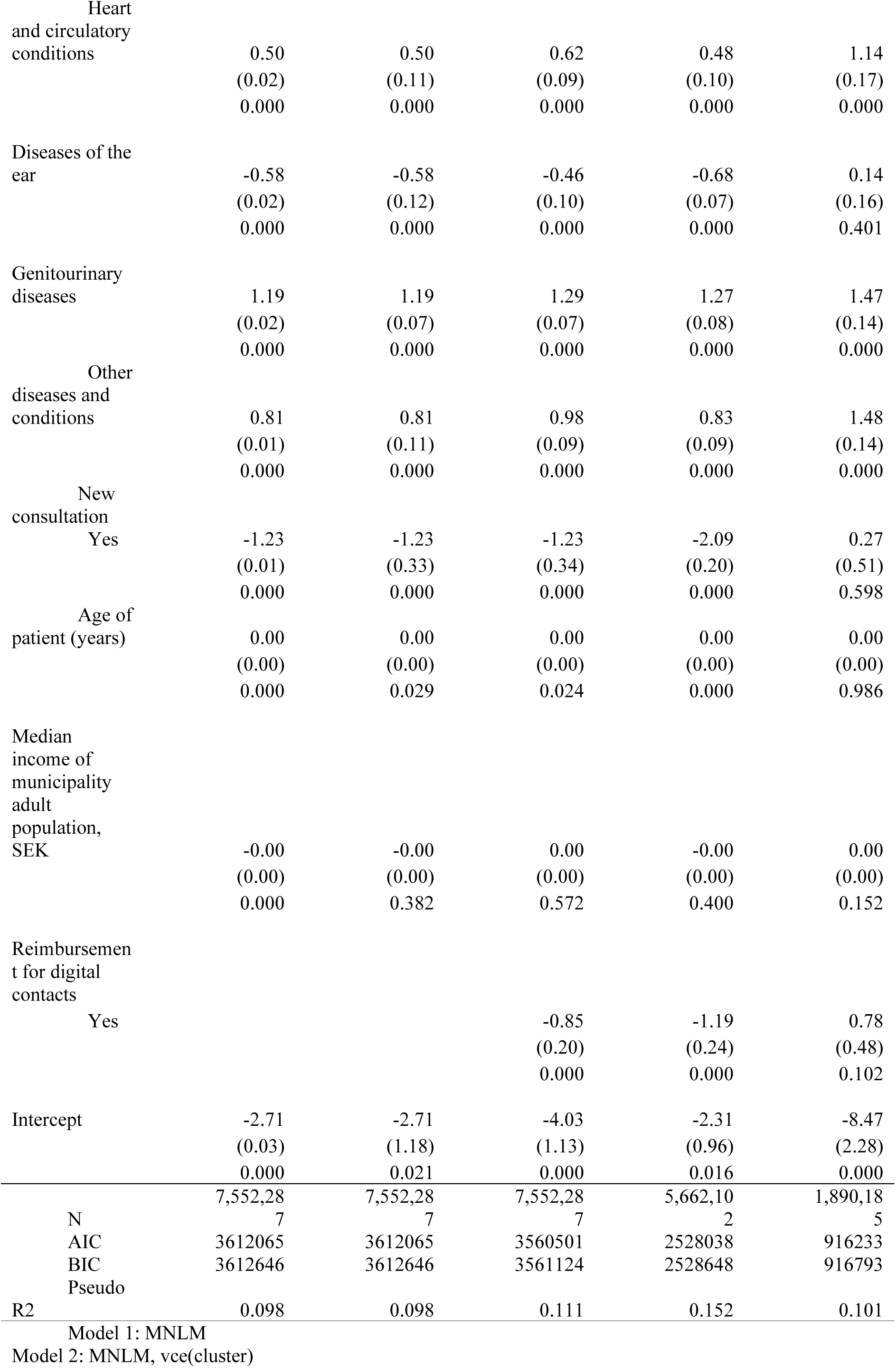
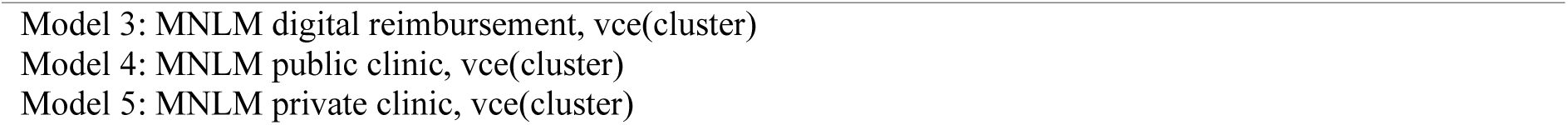
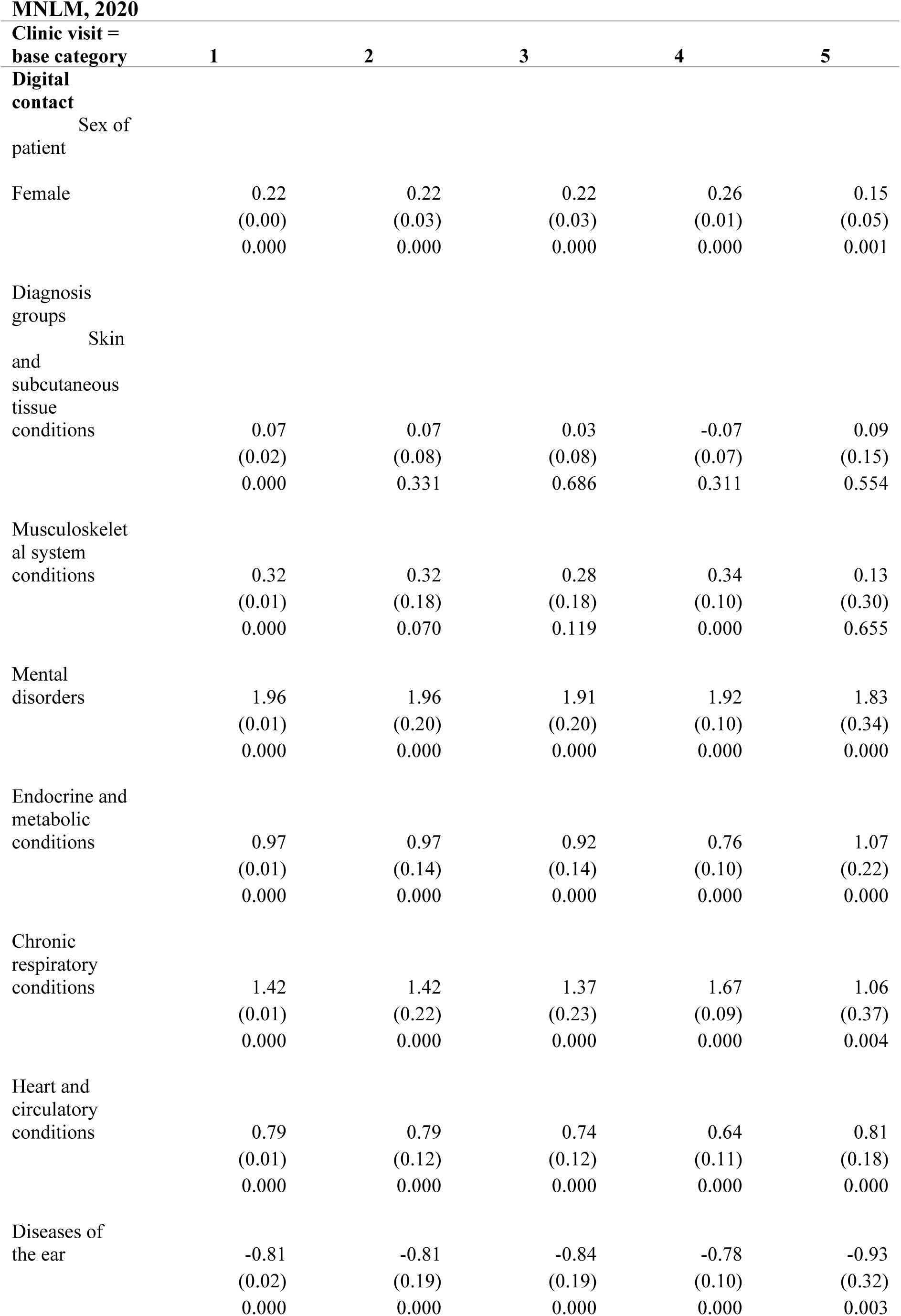
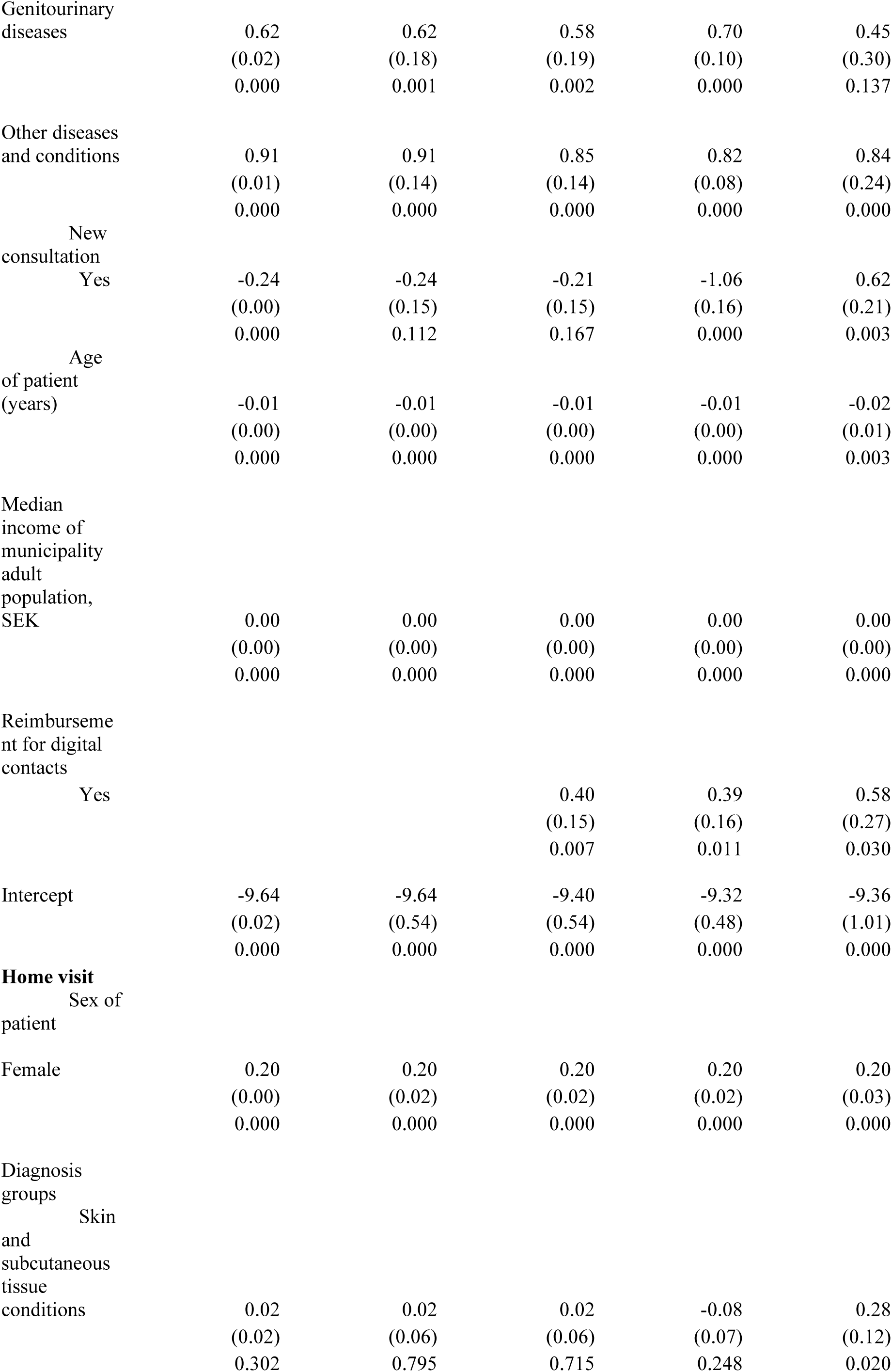
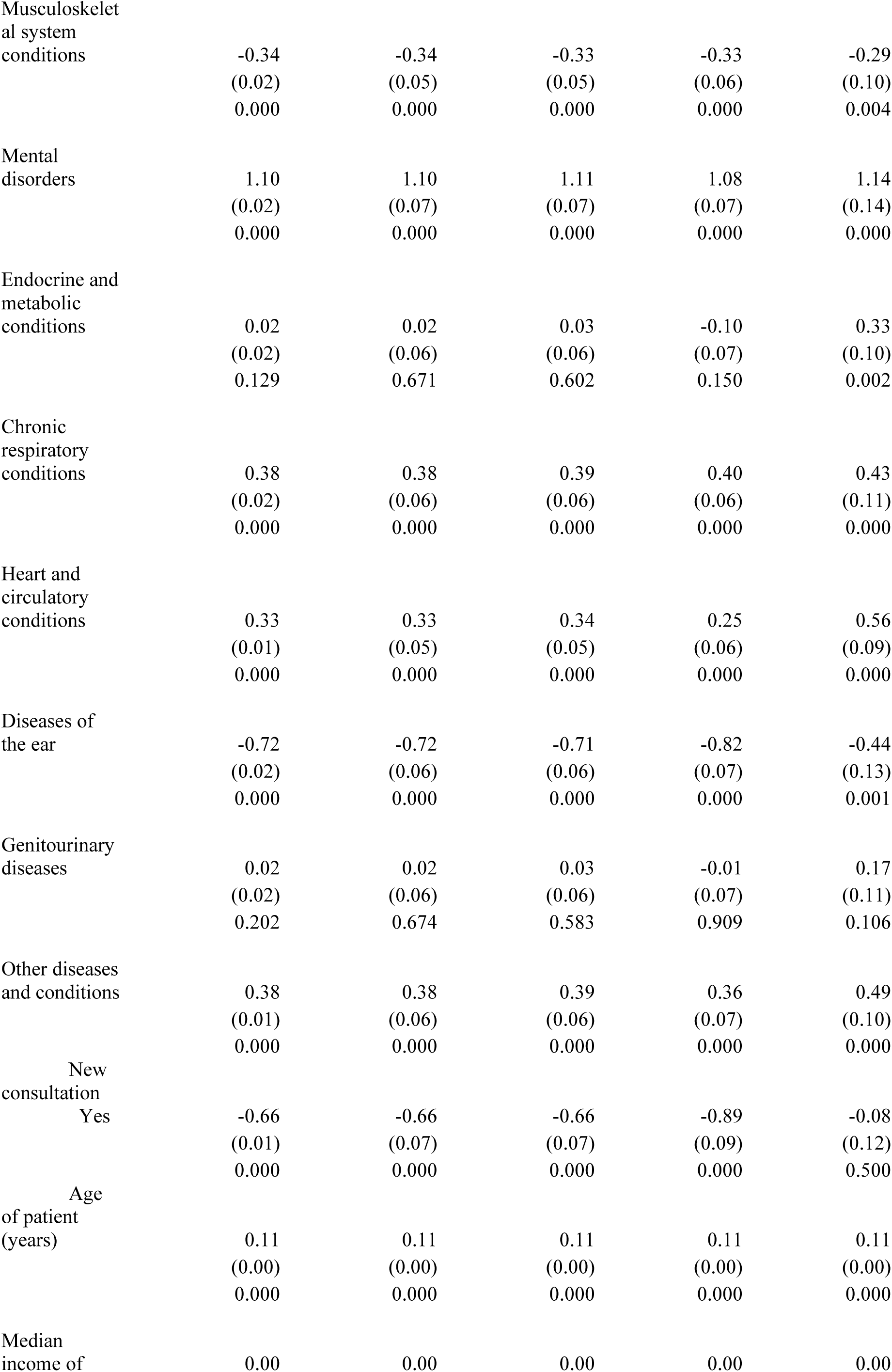
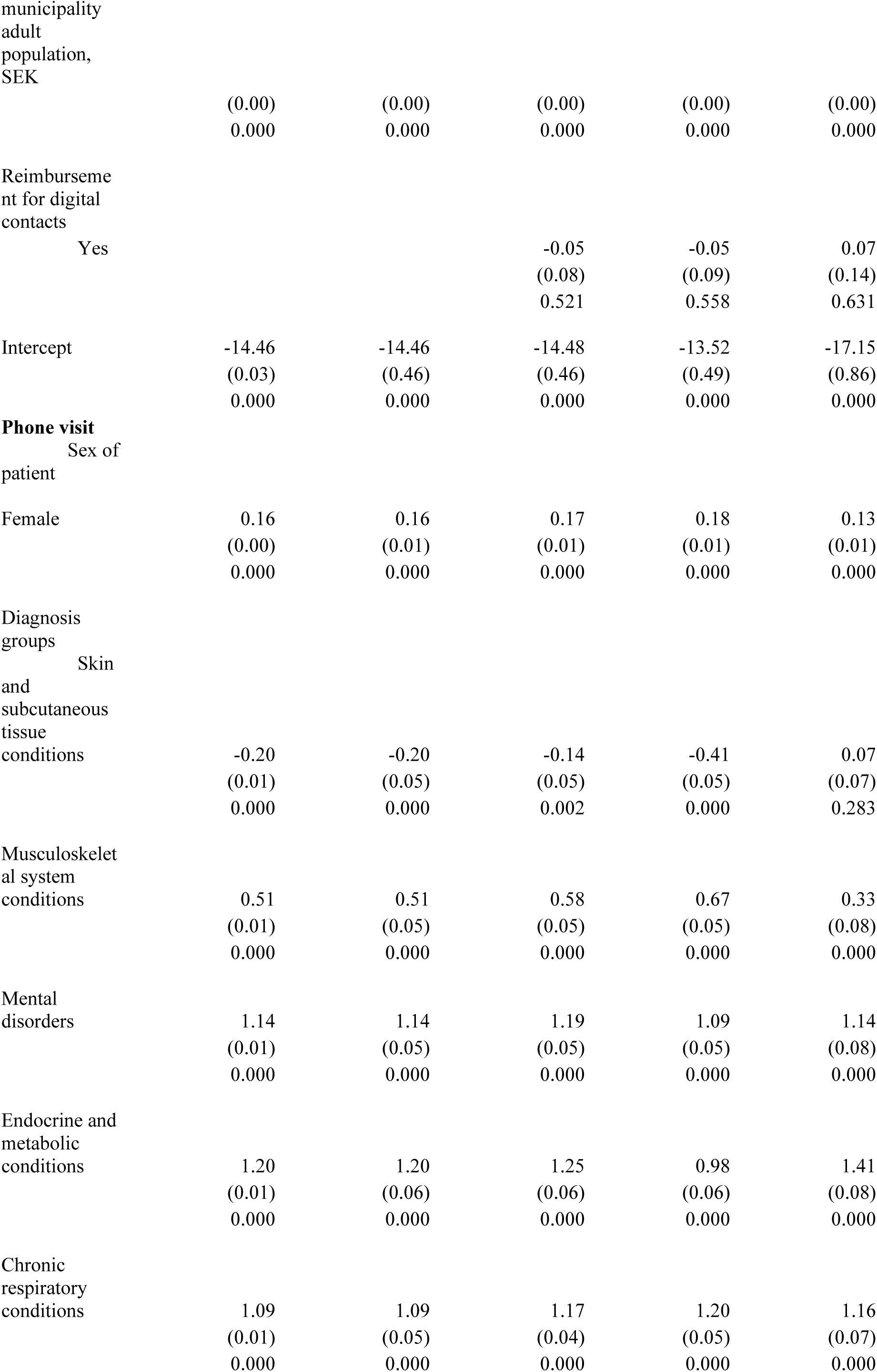
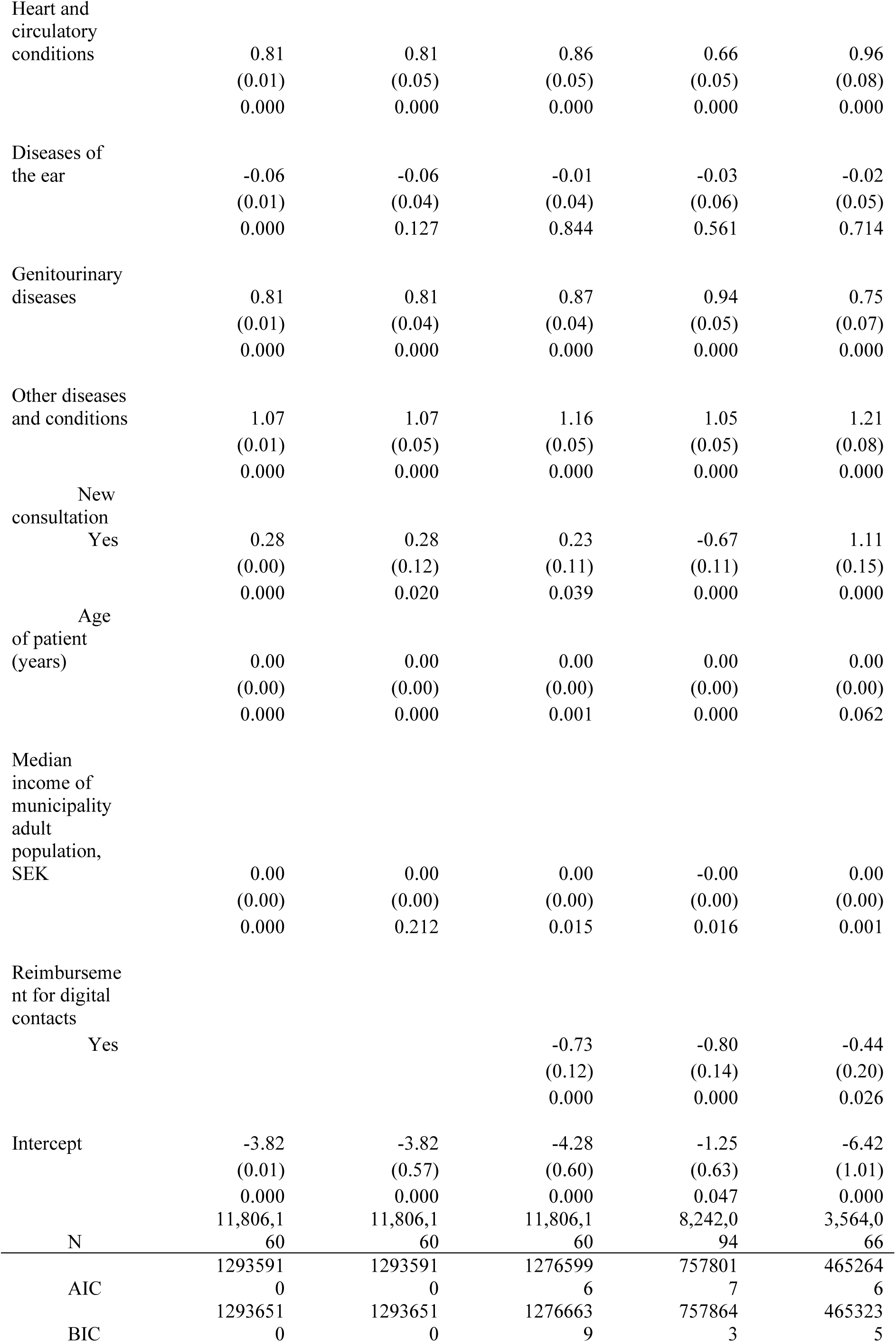
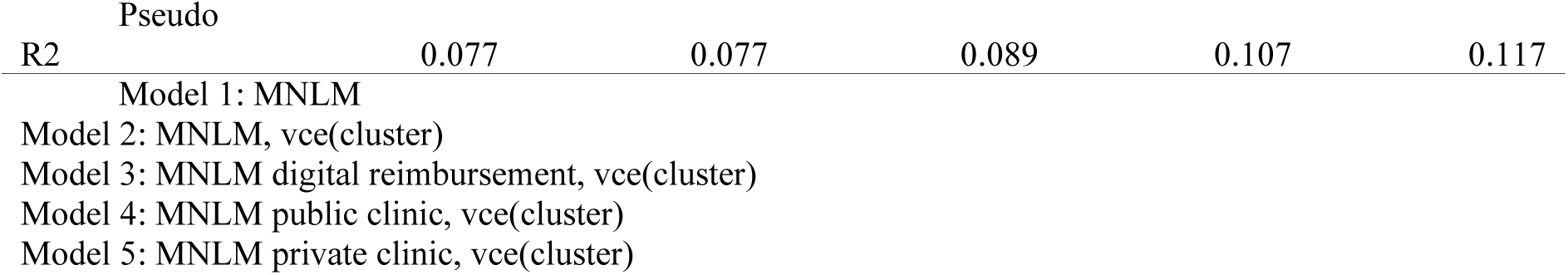
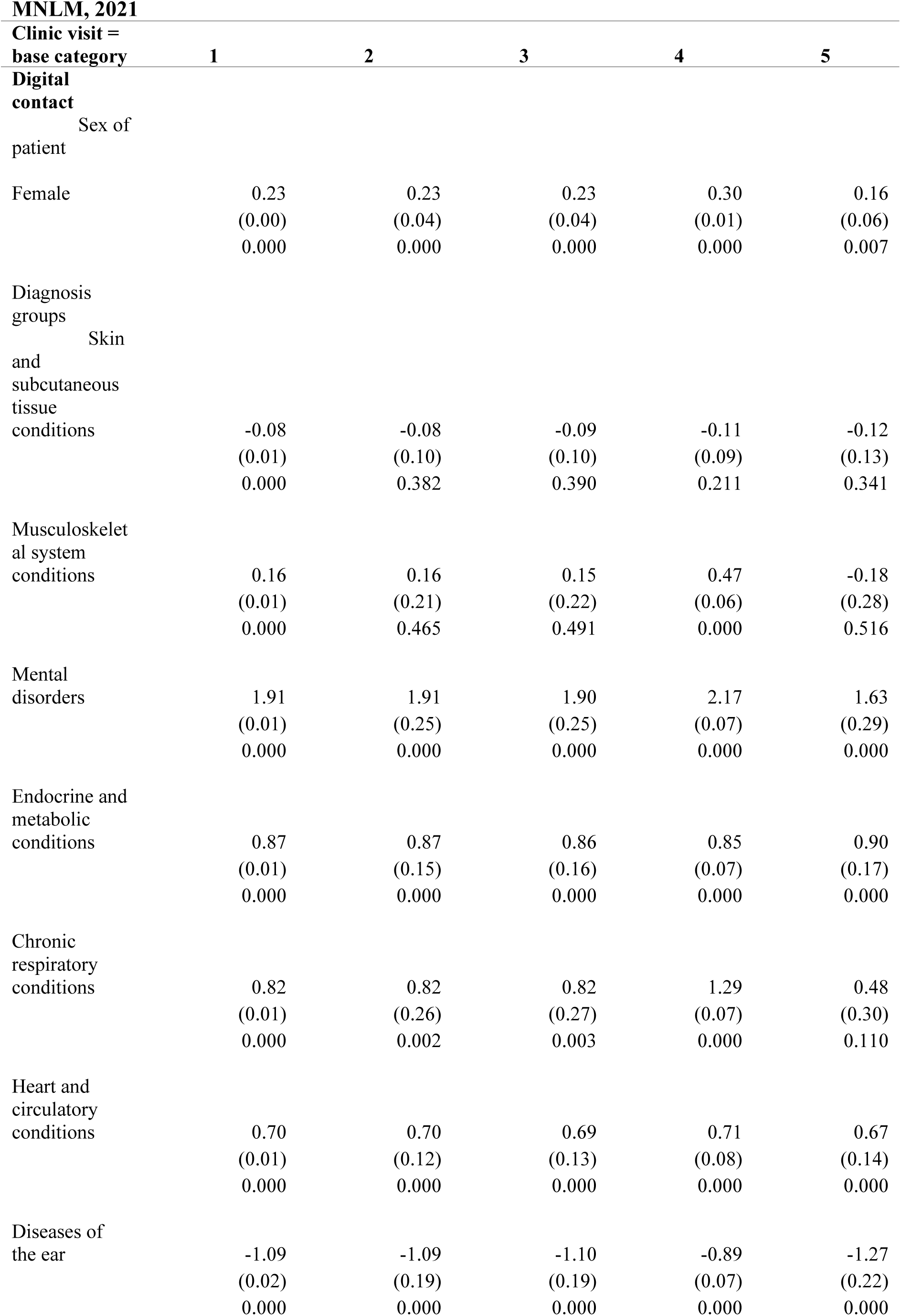
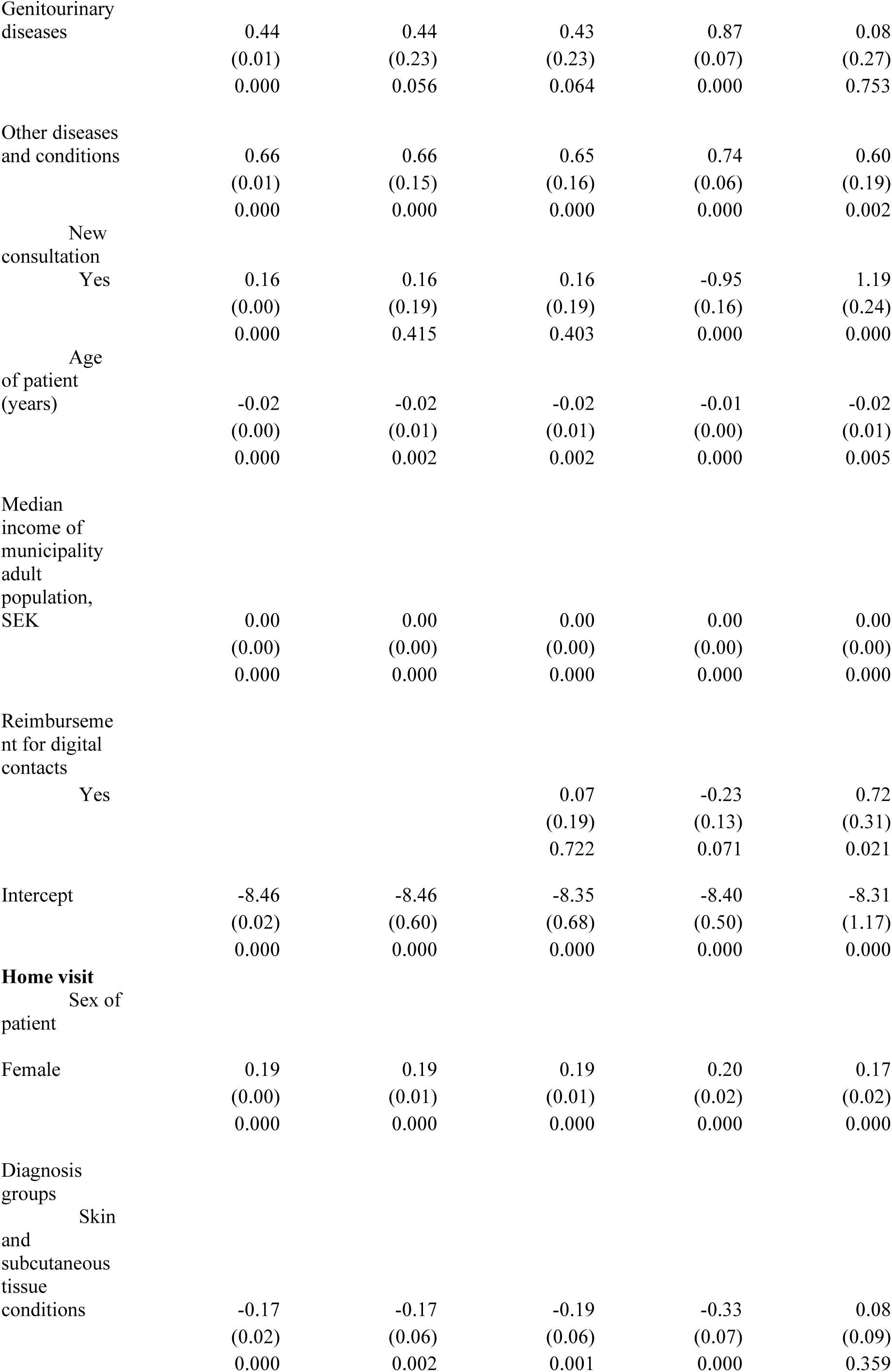
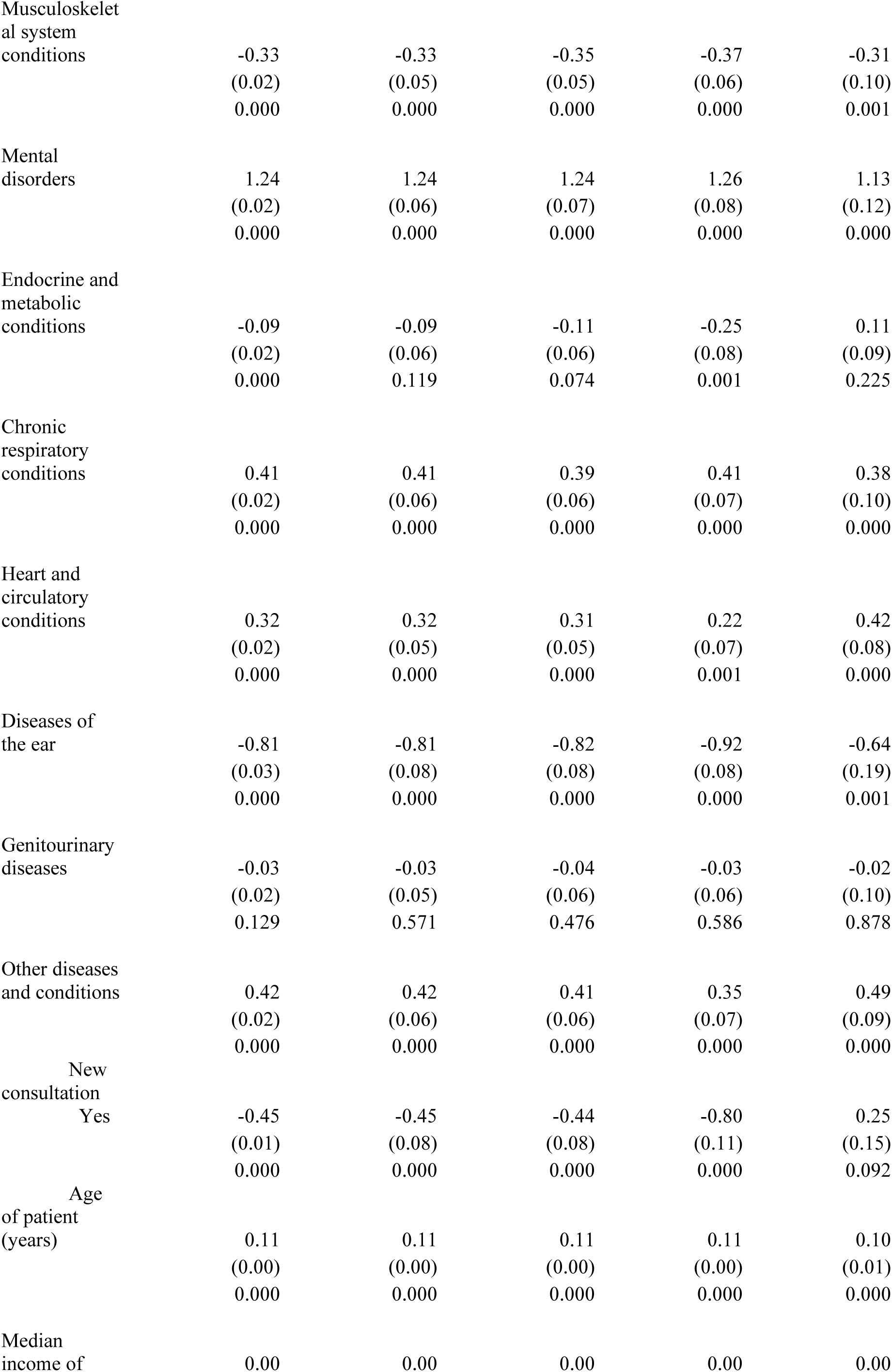
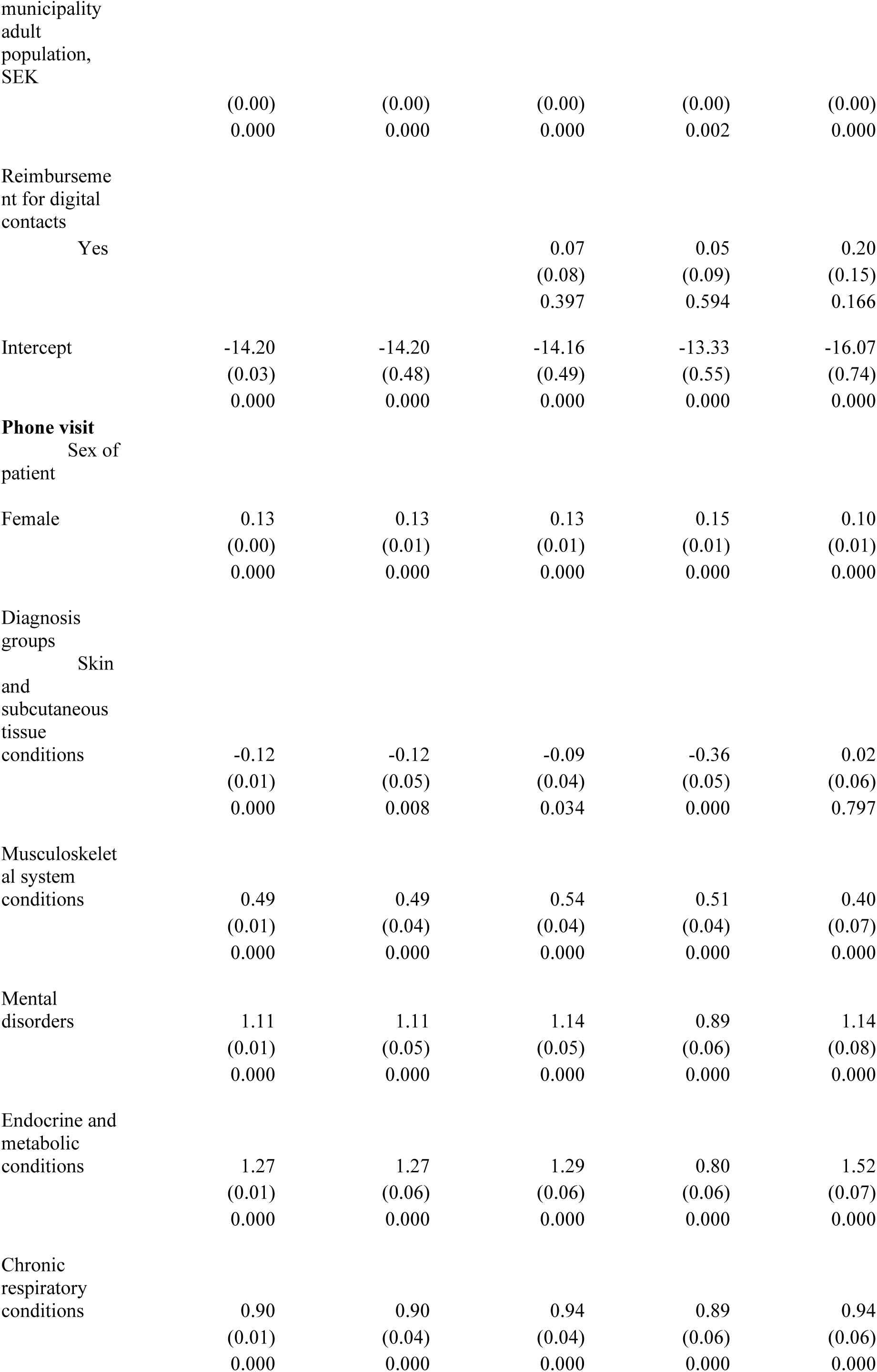
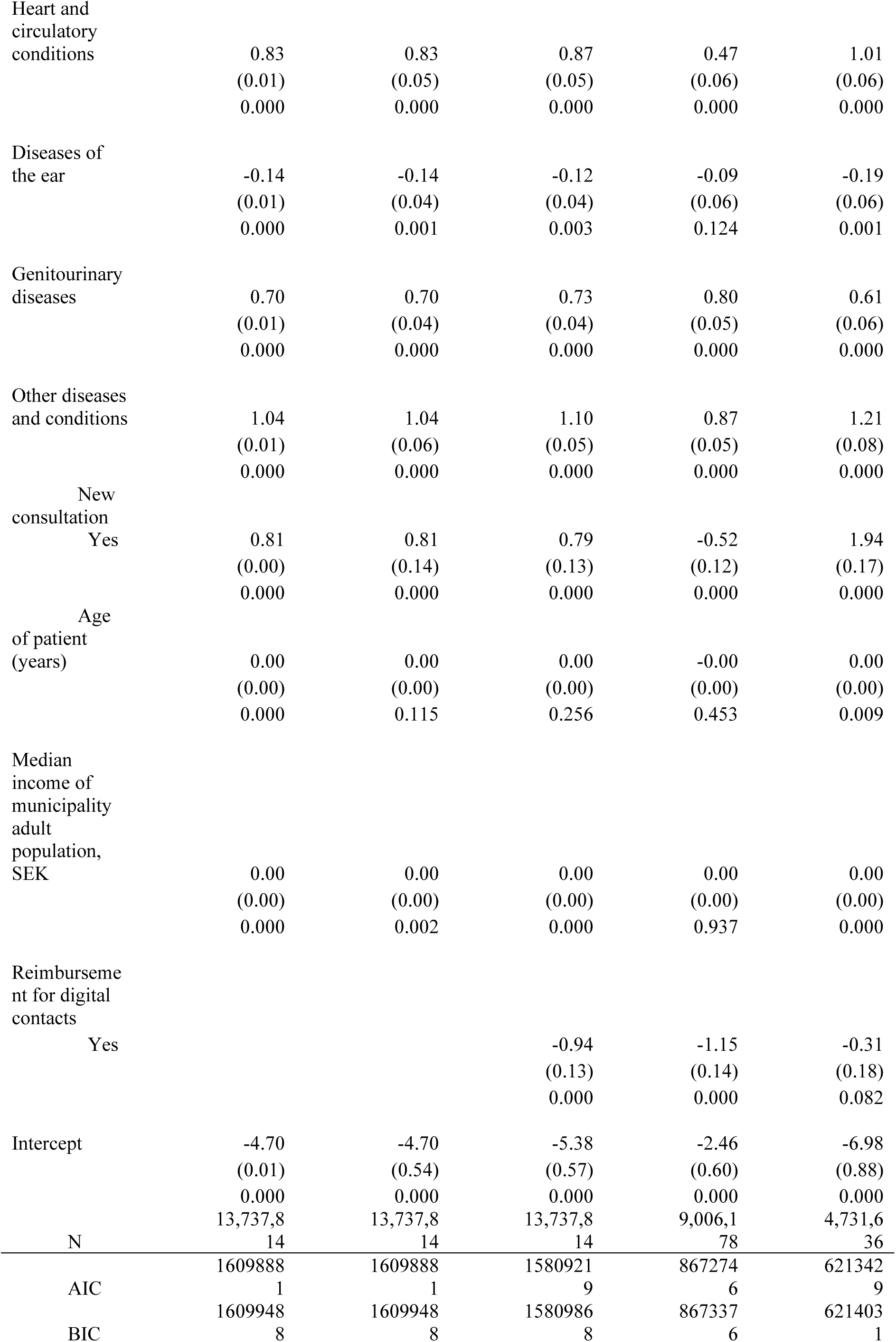
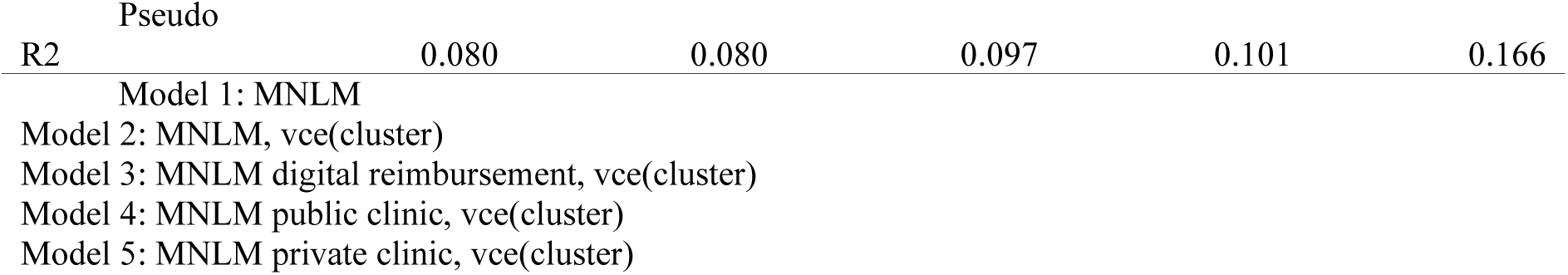
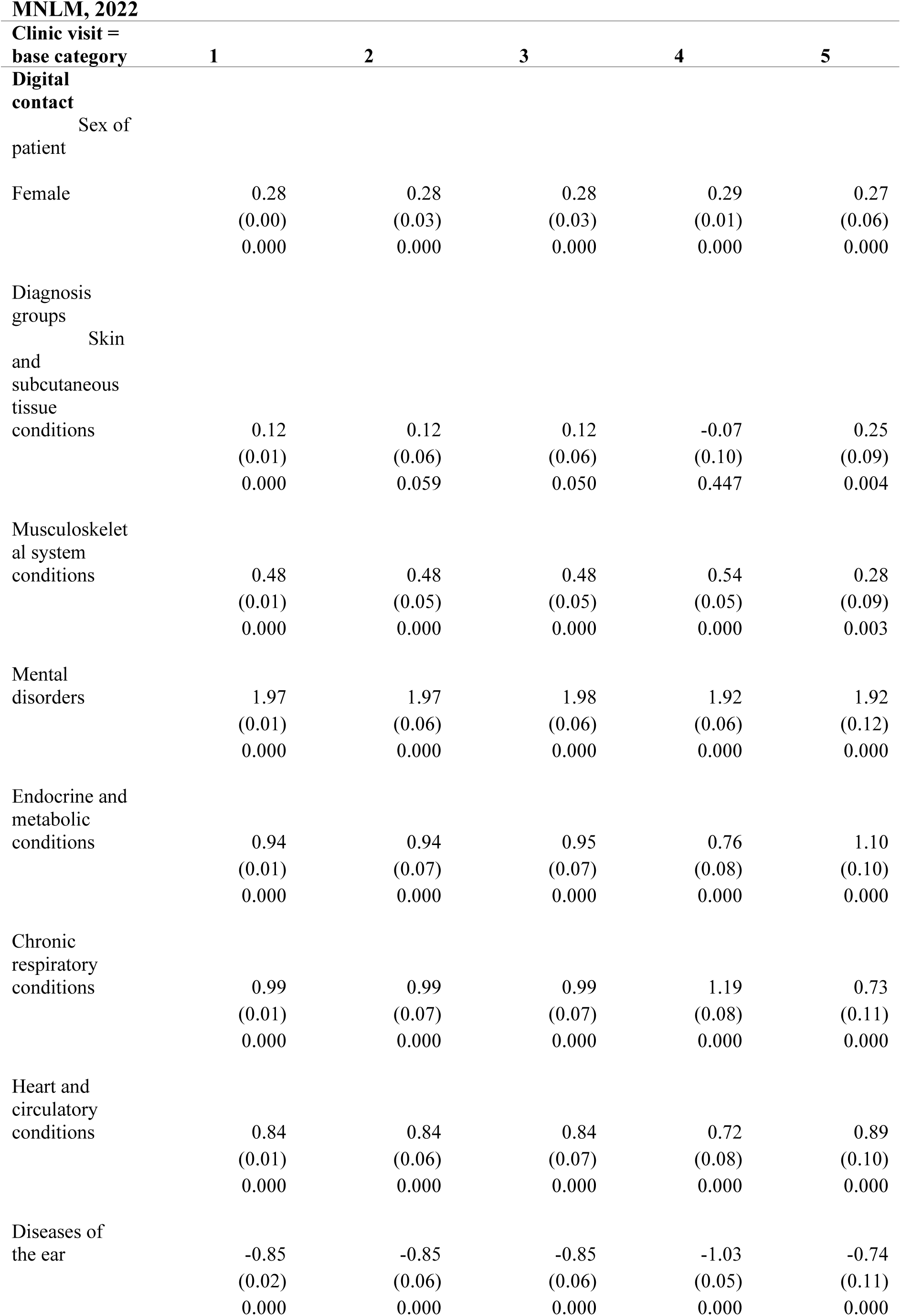
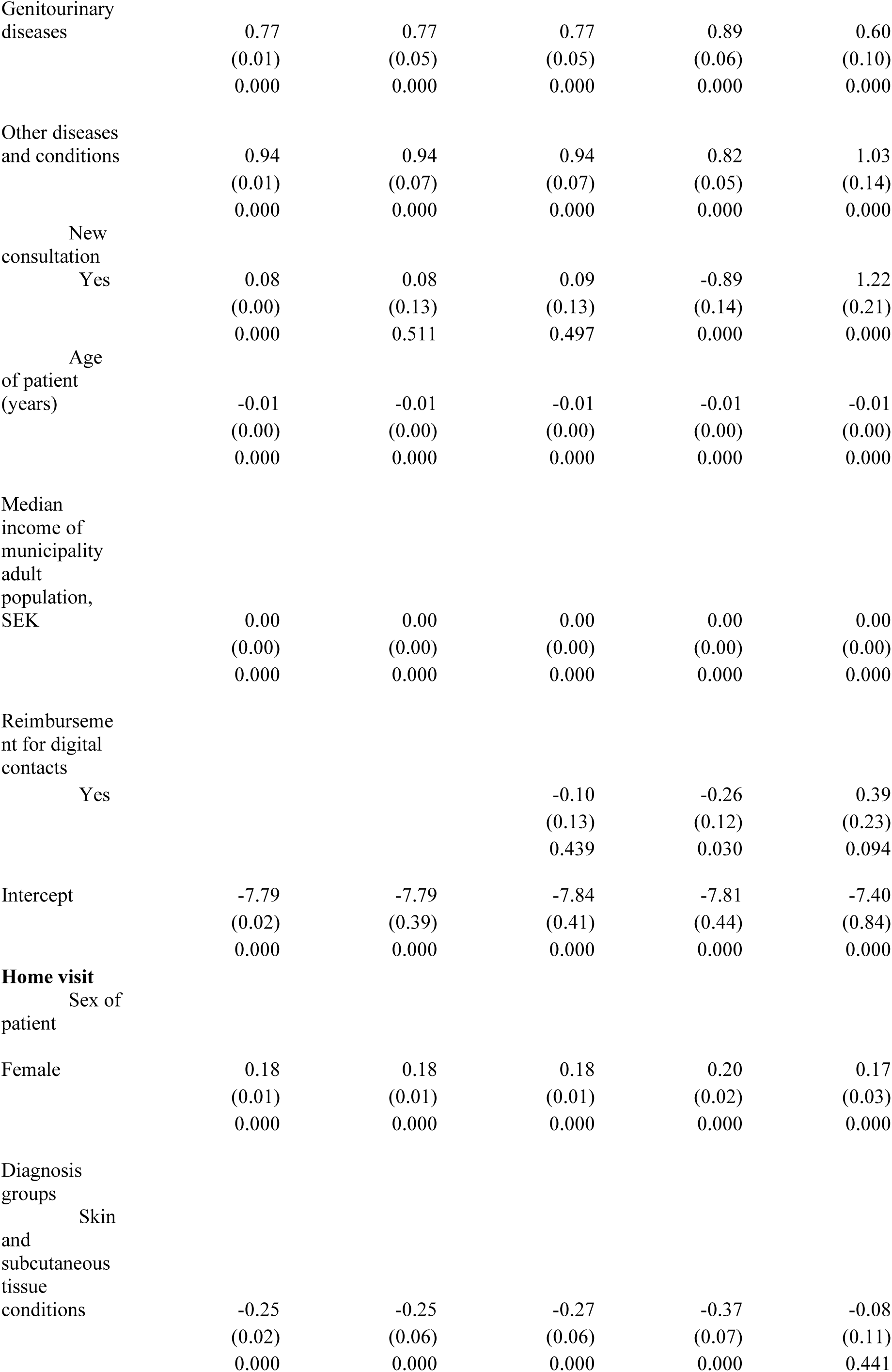
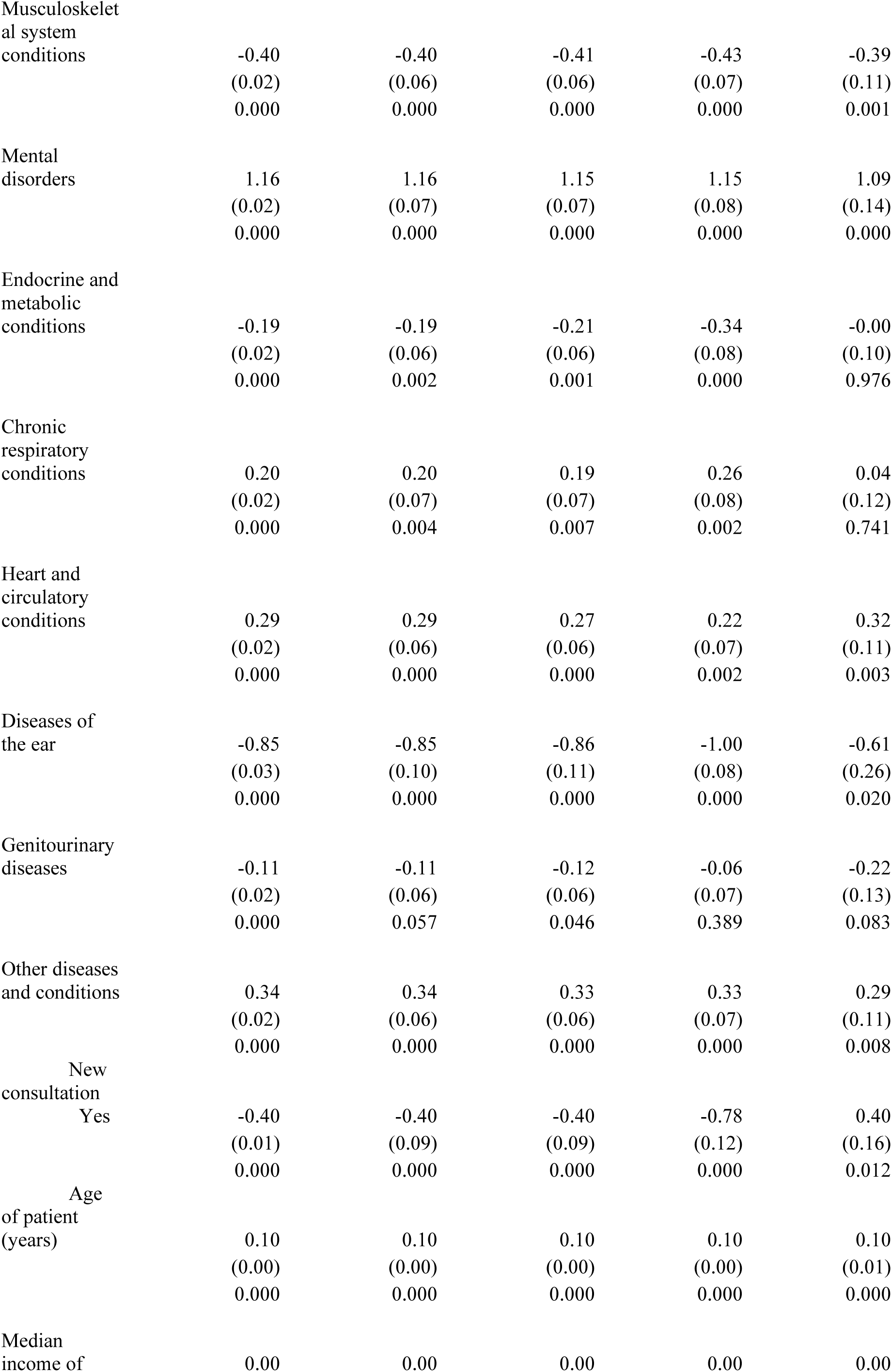
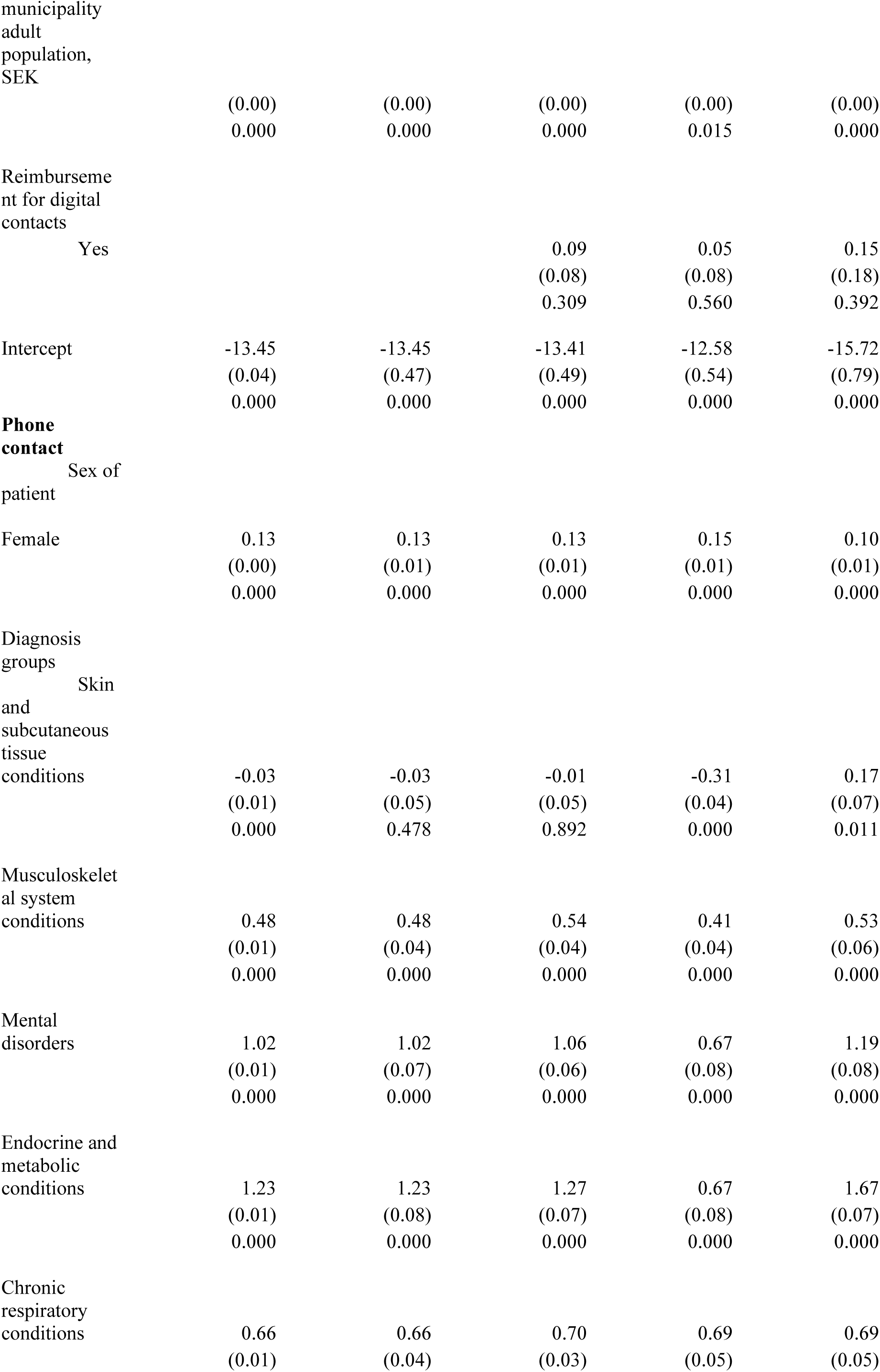
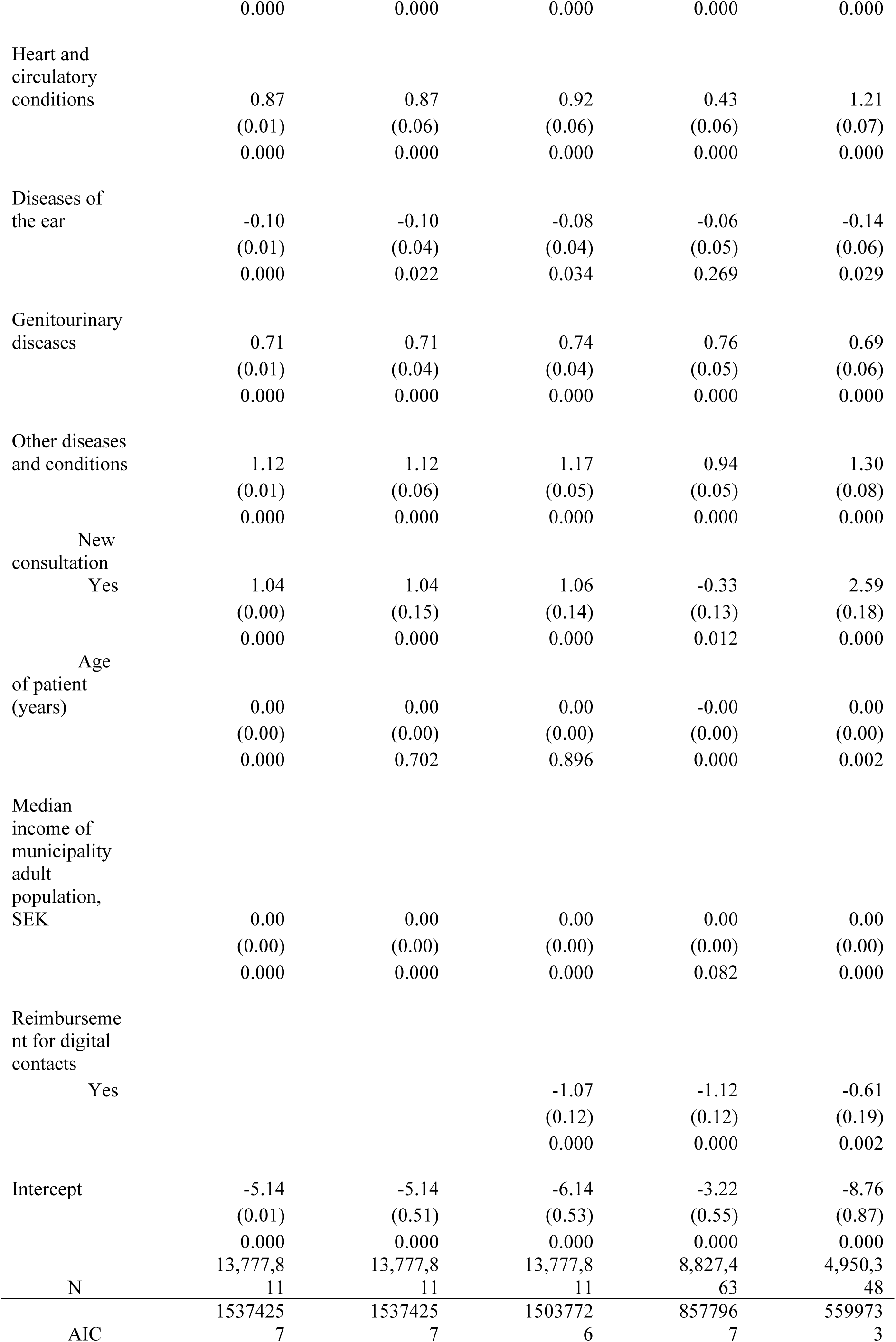
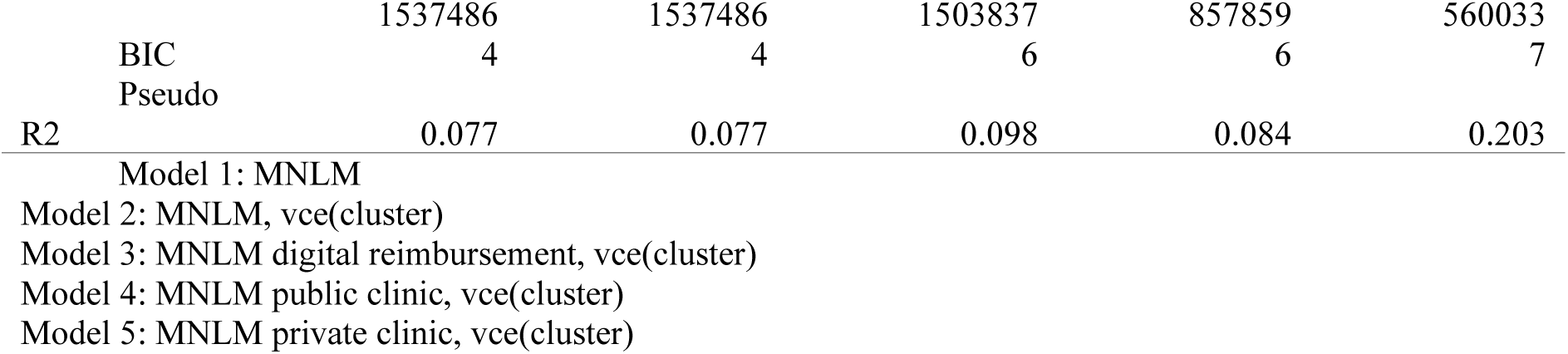

## Annex C. Graphs

**Figure C.1.**
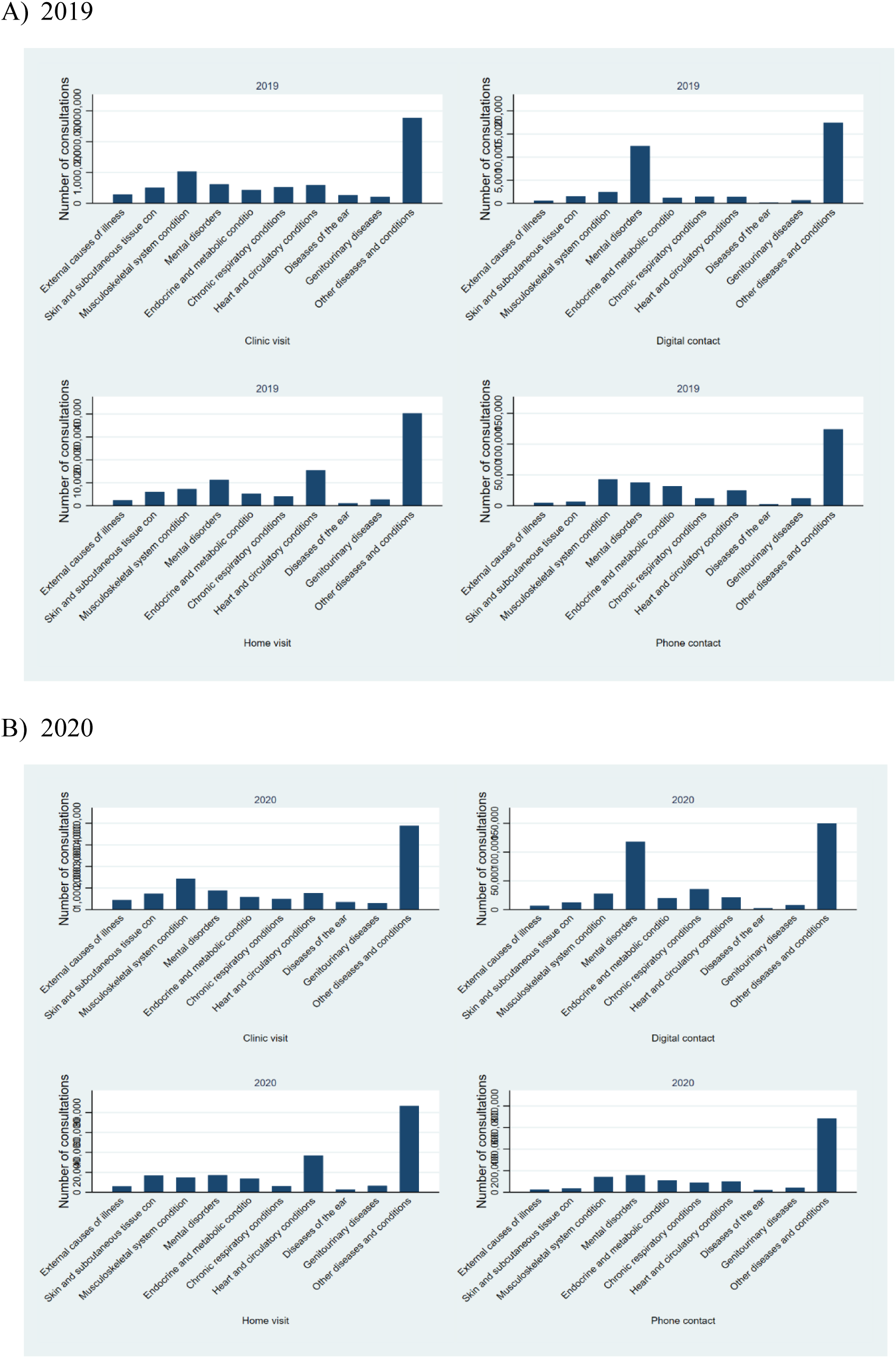
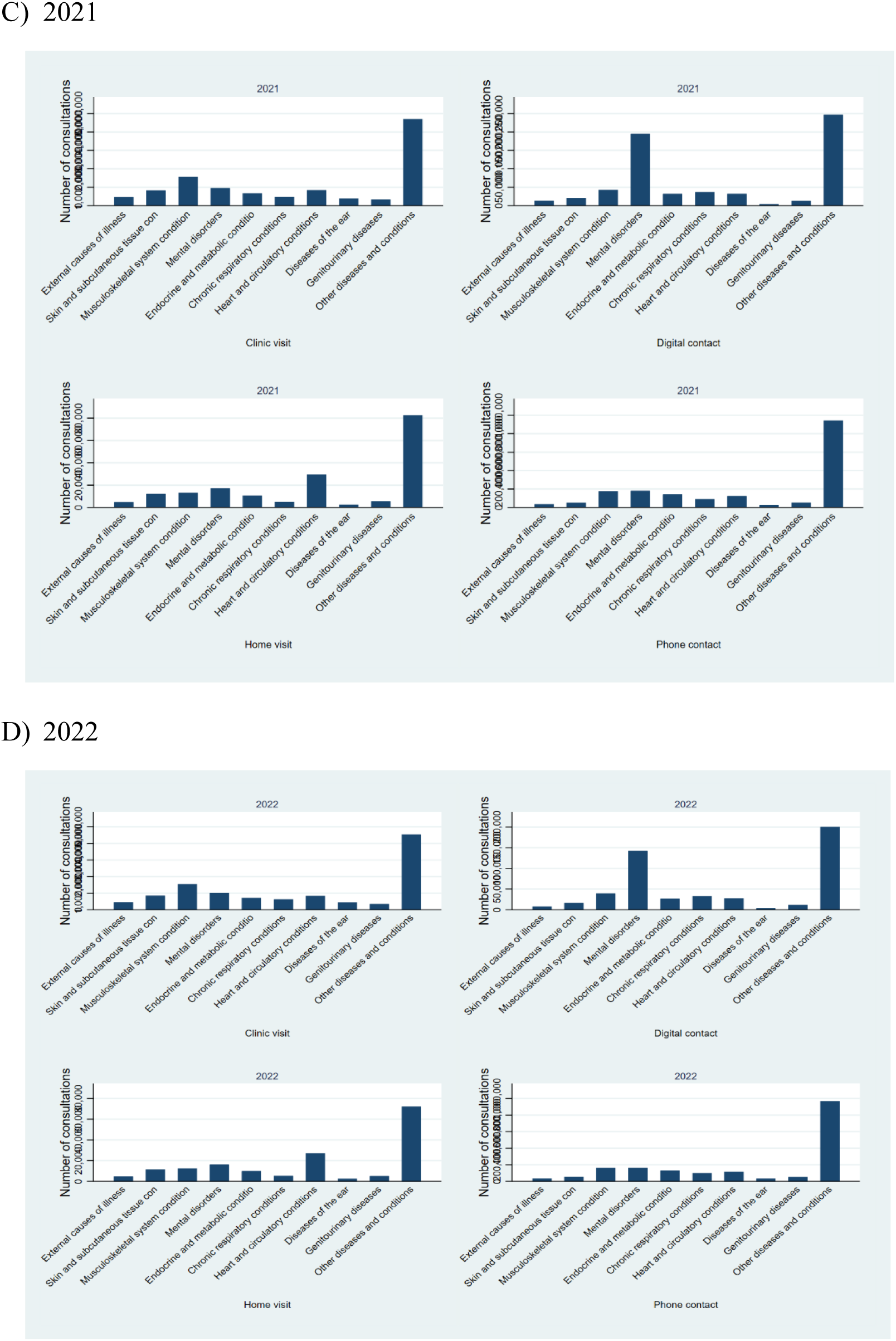
Type of consultation and diagnosis groups, 2019 – 2022.

**Figure C.2.**
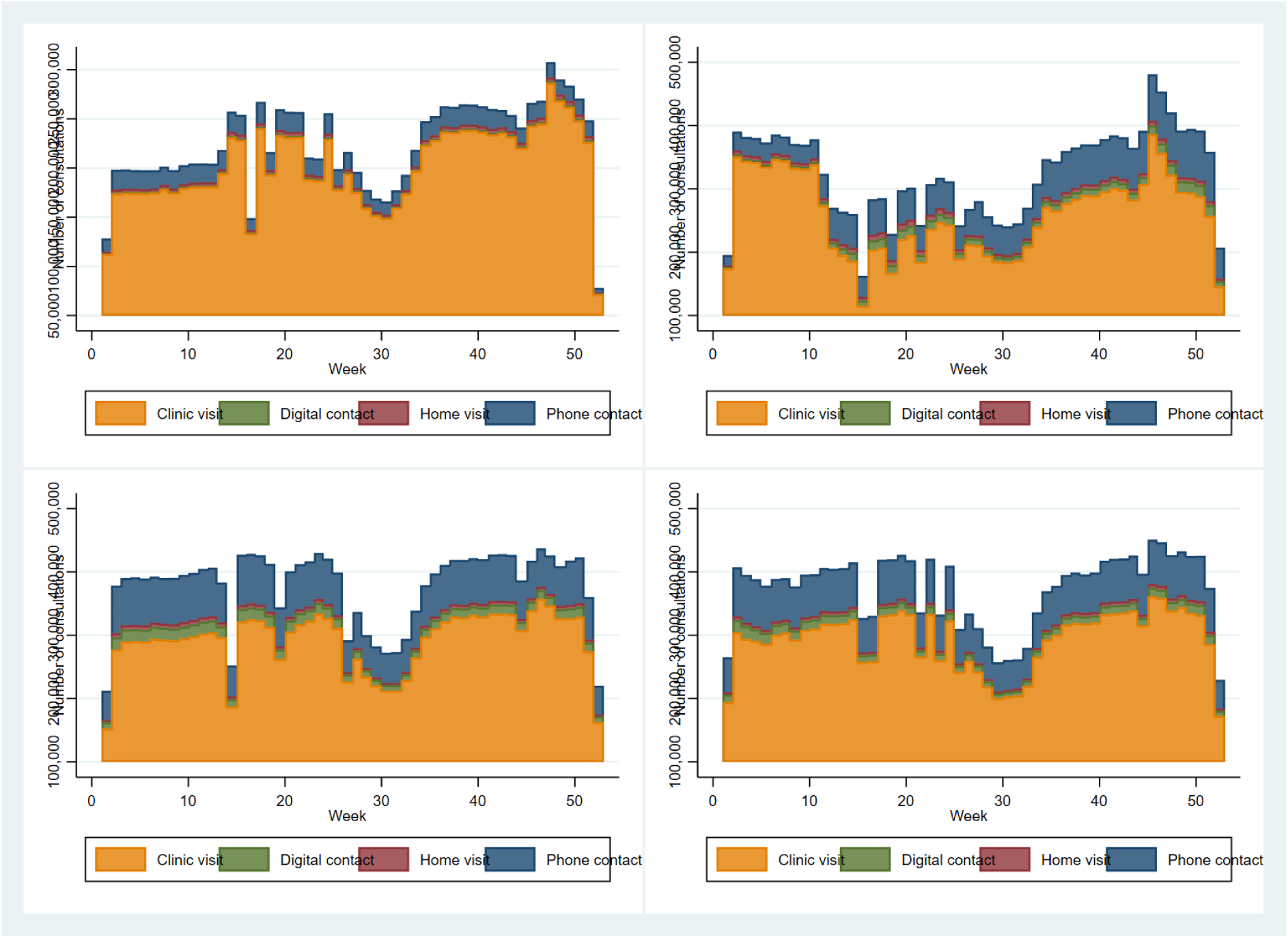
Type of consultation by week, 2019 – 2022.

**Figure C.3.**
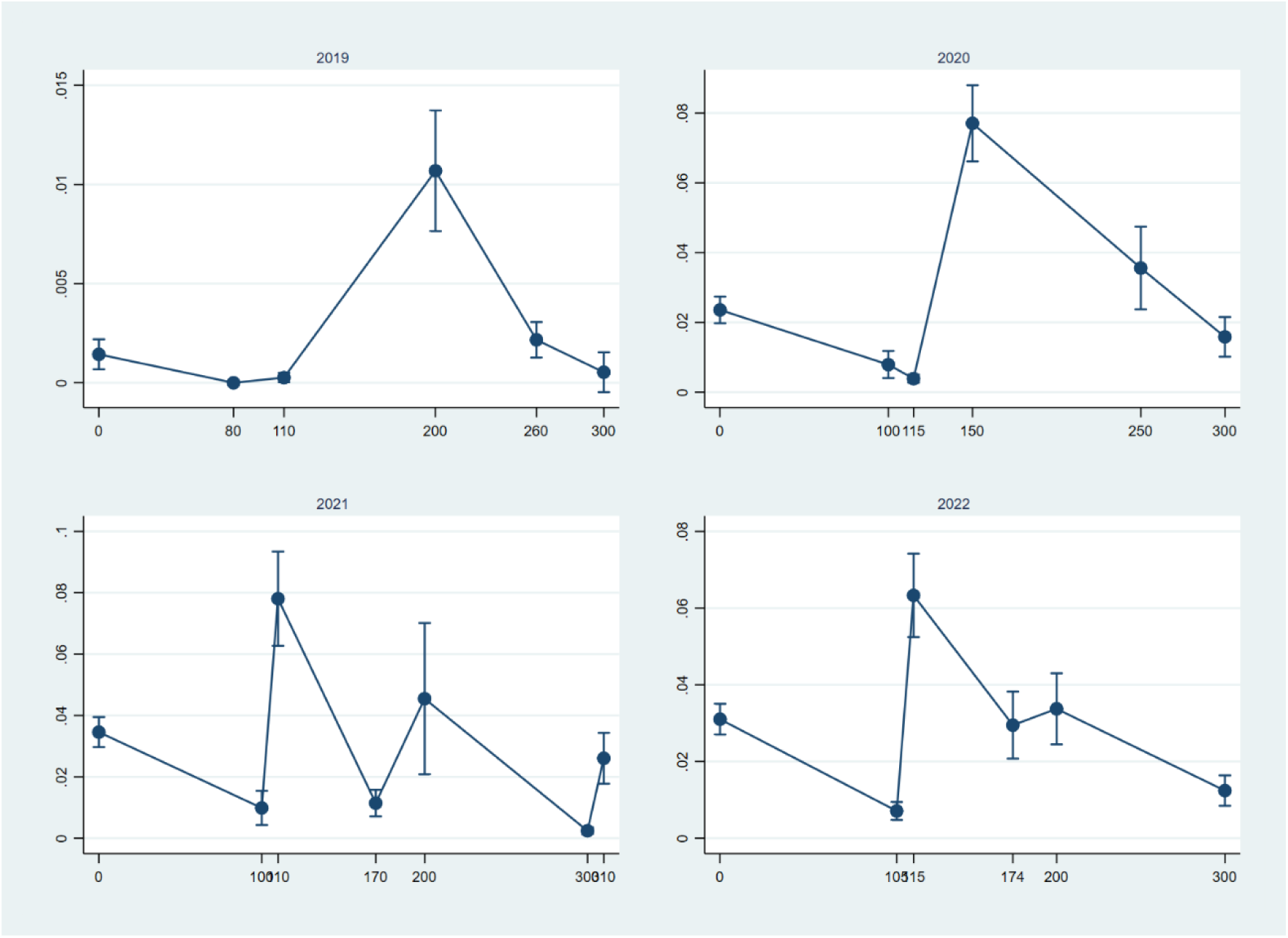
Predicted margins of digital reimbursement levels, 2019 – 2022.

## Notes

### Competing Interest Statement

The authors have declared no competing interest.

### Funding Statement

This study was funded by the Swedish Research Council.

## References

Altman, N. and M. Krzywinski (2018). "The curse(s) of dimensionality." Nature Methods 15(6): 399–400.

Blumenfeld, O. and J. Yaphe (2007). "What do smokers really fear? A survey of behavior, knowledge, and attitudes toward cigarette smoking among visitors to a primary care clinic in Israel." European Journal of General Practice 13(1): 40–41.

Cameron, A. C. and P. K. Trivedi (2022). Microeconometrics Using Stata. College Park, TX, Stata Press.

Cheung, L., T. I. Leung, V. Y. Ding, J. X. Wang, J. Norden, M. Desai, R. A. Harrington and S. Desai (2019). "Healthcare Service Utilization under a New Virtual Primary Care Delivery Model." Telemedicine Journal and e-Health 25(7): 551–559.

Clair, M. C. S., G. Sundberg and J. J. F. Kram (2019). "Incorporating Home Visits in a Primary Care Residency Clinic: The Patient and Physician Experience." Journal of Patient-Centered Research and Reviews 6(3): 203–209.

Ekman, B. (2017). "Cost Analysis of a Digital Health Care Model in Sweden." PhamacoEconomics Open 2(3): 347–354.

Ekman, B., E. Arvidsson, H. Thulesius, J. Wilkens and O. Cronberg (2021). "Impact of the Covid-19 pandemic on primary care utilization: evidence from Sweden using national register data." BMC Research Notes 14(424).

Fahy, N. and G. A. Williams (2021). Use of digital health tools in Europe: Before, during and after COVID-19. Policy Brief. Copenhagen, European Observatory on Health Systems and Policies.

Frees, E. W. (2004). Longitudinal and Panel Data: Analysis and Applications in the Social Sciences. Cambridge, UK, Cambridge University Press.

Grilli, G. and S. Ferrini (2022). "Discrete choice modeling in environmental and energy decision-making: an introduction to the special issue." Journal of Environmental Planning and Management 65(7): 1203–1209.

Kouskoukis, M.-N. and C. Botsaris (2017). "Cost-Benefit Analysis of Telemedicine Systems/Units in Greek Remote Areas." PharmacoEconomics - Open: Healthcare Interventions and Outcomes 1(2): 117–121.

Lake, R., A. Georgiou, J. Li, L. Li, M. Byrne, M. Robinson and J. I. Westbrook (2017). "The quality, safety and governance of telephone triage and advice services – an overview of evidence from systematic reviews." BMC Health Services Research 17(1): 614.

Long, J. S. and J. Freese (2014). Regression Models for Categorical Dependent Variables Using Stata. College Station, Stata Press.

Marcinowicz, L., S. Chlabicz and Z. Gugnowski (2007). "Home visits by family physicians in Poland: Patients’ perspective." European Journal of General Practice 13(4): 237–238.

Neves, A. L. and J. Burgers (2022). "Digital technologies in primary care: Implications for patient care and future research." European Journal of General Practice 28(1): 203–208.

Odgers, C. L. and M. R. Jensen (2020). "Adolescent development and growing divides in the digital age." Dialogues in Clinical Neuroscience 22(2): 143–149.

Piera-Jiménez, J., T. Dedeu, C. Pagliari and T. Trupec (2024). "Strengthening primary health care in Europe with digital solutions." Atención Primaria 56(10): 102904.

Rabe-Hesketh, S. and A. Skrondal (2012). Multilevel and Longitudinal Modeling Using Stata. College Park, TX, Stata Press.

Ritchie, H., E. Mathiew, M. Roser and E. Ortiz-Ospina (2023). Internet, Our World in Data.

StataCorp (2021). Choice Models Reference Manual, Release 18. College Park, TA, Stata Press.

StataCorp (2021). Structural Equation Modeling Reference Manual, Release 17. Statistical Software. College Station, TX, StataCorp LLC.

Stoffers, J. (2018). "The promise of eHealth for primary care: opportunities for service delivery, patient–doctor communication, self-management, shared decision making and research." European Journal of General Practice 24(1): 146–148.

Sun, C.-A., C. Parslow, J. L. Gray, I. Koyfman, M. d. Hladek and H.-R. Han (2022). "Home-based primary care visits by nurse practitioners." Journal of the American Association of Nurse Practitioners 34(6).

Toal-Sullivan, D., S. Dahrouge, J. Tesfaselassie and L. Olejnik (2024). "Access to primary health care: perspectives of primary care physicians and community stakeholders." BMC Primary Care 25(1): 1–12.

Torre-Diez, I. d., M. López-Coronado, C. Vaca, J. S. Aguado and C. d. Castro (2015). "Cost-Utility and Cost-Effectiveness Studies of Telemedicine, Electronic, and Mobile Health Systems in the Literature: A Systematic Review." Telemedicin and e-Health 21(2): 81–85.

van der Kleij, R. M. J. J., M. J. Kasteleyn, E. Meijer, T. N. Bonten, E. J. F. Houwink, M. Teichert, S. van Luenen, R. Vedanthan, A. Evers, J. Car, H. Pinnock and N. H. Chavannes (2019). "SERIES: eHealth in primary care. Part 1: Concepts, conditions and challenges." European Journal of General Practice 25(4): 179–189.

Young, H., T. Nesbitt, H. M. Young and T. S. Nesbitt (2017). "Increasing the Capacity of Primary Care Through Enabling Technology." JGIM: Journal of General Internal Medicine 32(4): 398–403.

Zanaboni, P., P. Nguangue, G. I. C. Mbemba, T. R. Schopf, T. S. Bergmo and M.-P. Gagnon (2018). "Methods to Evaluate the Effect of Internet-Based Digital Health Interventions for Citizens: Systematic Reivew of Reviews." Journal of Medical Internet Research 20(6): e10202.

